# Multicenter analysis of neutrophil extracellular trap dysregulation in adult and pediatric COVID-19

**DOI:** 10.1101/2022.02.24.22271475

**Authors:** Carmelo Carmona-Rivera, Yu Zhang, Kerry Dobbs, Tovah E. Markowitz, Clifton L. Dalgard, Andrew J. Oler, Dillon R. Claybaugh, Deborah Draper, Meng Truong, Ottavia M. Delmonte, Francesco Licciardi, Ugo Ramenghi, Nicoletta Crescenzio, Luisa Imberti, Alessandra Sottini, Virginia Quaresima, Chiara Fiorini, Valentina Discepolo, Andrea Lo Vecchio, Alfredo Guarino, Luca Pierri, Andrea Catzola, Andrea Biondi, Paolo Bonfanti, Maria Cecilia Poli Harlowe, Yasmin Espinosa, Camila Astudillo, Emma Rey-Jurado, Cecilia Vial, Javiera de la Cruz, Ricardo Gonzalez, Cecilia Pinera, Jacqueline W. Mays, Ashley Ng, Andrew Platt, NIH COVID Autopsy Consortium, COVID STORM Clinicians, Beth Drolet, John Moon, Edward W. Cowen, Heather Kenney, Sarah E. Weber, Riccardo Castagnoli, Mary Magliocco, Michael A. Stack, Gina Montealegre, Karyl Barron, Stephen M. Hewitt, Lisa M. Arkin, Daniel S. Chertow, Helen C. Su, Luigi D. Notarangelo, Mariana J. Kaplan

**Affiliations:** Systemic Autoimmunity Branch, National Institute of Arthritis and Musculoskeletal and Skin Diseases (NIAMS), National Institutes of Health (NIH), Bethesda, MD, USA; Human Immunological Diseases Section, Laboratory of Clinical Immunology and Microbiology (LCIM), National Institute of Allergy and Infectious Diseases (NIAID), NIH, Bethesda, MD, USA; LCIM, NIH, Bethesda, MD, USA; Bioinformatics and Computational Biosciences Branch, Office of Cyber Infrastructure and Computational Biology, NIAID, NIH, Bethesda, MD; Axle Informatics, Bethesda, MD, USA; Department of Anatomy, Physiology & Genetics, School of Medicine, Uniformed Services University of the Health Sciences (USUHS), Bethesda, MD and The American Genome Center, USUHS, Bethesda, MD, USA; Department of Public Health and Pediatric Sciences, “Regina Margherita” Children’s Hospital, University of Turin, Turin, Italy; Pediatric Hematology, “Regina Margherita” Children Hospital, University of Turin, Turin, Italy; Centro di Ricerca Emato-oncologica AIL (CREA), Diagnostic Department, ASST Spedali Civili di Brescia, Brescia, Italy; Department of Translational Medical Sciences, Pediatric Section, University of Naples Federico II, Naples, Italy; Department of Pediatrics, University of Milano-Bicocca, European Reference Network (ERN) PaedCan, EuroBloodNet, MetabERN, Fondazione MBBM/Ospedale San Gerardo, Monza, Italy; Department of Infectious Diseases, San Gerardo Hospital–University of Milano-Bicocca, Monza, Italy; Programa de Inmunogenética e Inmunología Traslacional, Instituto de Ciencias e Innovación en Medicina, Facultad de Medicina Clínica Alemana, Universidad del Desarrollo, Santiago, Chile; Hospital Roberto del Rio, Santiago, Chile; Facultad de Medicina Clínica Alemana Universidad del Desarrollo, Programa Hantavirus, Instituto de Ciencias e Innovación en Medicina, Santiago, Chile; Pediatric Intensive Care Unit, Hospital Exequiel Gonzalez Cortés, Santiago, Chile; Infectious Diseases Unit, Hospital Dr. Exequiel González Cortés, Región Metropolitana, Chile; Faculty of Medicine, Universidad de Chile, Santiago, Chile; National Institute of Dental and Craniofacial Research, NIH, Bethesda, MD, USA; Department of Dermatology, University of Wisconsin School of Medicine and Public Health, Madison, WI, USA; Emerging Pathogens Section, Critical Care Medicine Department, Clinical Center, and Laboratory of Immunoregulation, NIAID, NIH, Bethesda, MD, USA; Molecular Development of the Immune System Section, Laboratory of Immune System Biology, NIAID, NIH, Bethesda, MD; Dermatology Branch, NIAMS, NIH, Bethesda, MD, USA; Division of Clinical Research, NIAID, NIH, Bethesda, MD; Laboratory of Pathology, Center for Cancer Research, National Cancer Institute, NIH, Bethesda, MD, USA

## Abstract

Dysregulation in neutrophil extracellular trap (NET) formation and degradation may play a role in the pathogenesis and severity of COVID-19; however, its role in the pediatric manifestations of this disease including MIS-C and chilblain-like lesions (CLL), otherwise known as “COVID toes”, remains unclear. Studying multinational cohorts, we found that, in CLL, NETs were significantly increased in serum and skin. There was geographic variability in the prevalence of increased NETs in MIS-C, in association with disease severity. MIS-C and CLL serum samples displayed decreased NET degradation ability, in association with C1q and G-actin or anti-NET antibodies, respectively, but not with genetic variants of DNases. In adult COVID-19, persistent elevations in NETs post-disease diagnosis were detected but did not occur in asymptomatic infection. COVID-19-affected adults displayed significant prevalence of impaired NET degradation, in association with anti-DNase1L3, G-actin, and specific disease manifestations, but not with genetic variants of DNases. NETs were detected in many organs of adult patients who died from COVID-19 complications. Infection with the Omicron variant was associated with decreased levels of NETs when compared to other SARS-CoV-2 strains. These data support a role for NETs in the pathogenesis and severity of COVID-19 in pediatric and adult patients.

**Summary:** NET formation and degradation are dysregulated in pediatric and symptomatic adult patients with various complications of COVID-19, in association with disease severity. NET degradation impairments are multifactorial and associated with natural inhibitors of DNase 1, G-actin and anti-DNase1L3 and anti-NET antibodies. Infection with the Omicron variant is associated with decreased levels of NETs when compared to other SARS-CoV-2 strains.

## Introduction

Coronavirus disease 2019 (COVID-19) is caused by infection with severe acute respiratory syndrome coronavirus 2 (SARS-CoV-2) (Atzrodt et al., 2020; Li et al., 2020). SARS-CoV-2 infection causes mild to severe illness that may progress to acute respiratory distress syndrome, multiorgan failure, and death. In severe cases, widespread infection of pneumocytes and associated inflammation results in lung injury, thrombotic microangiopathy and hypoxemia (Azkur et al., 2020; Costela-Ruiz et al., 2020; Diorio et al., 2020; Hu et al., 2021; Miesbach and Makris, 2020; Tao et al., 2021; Vinayagam and Sattu, 2020). More than 5 million COVID-19 deaths have been reported globally, predominantly among unvaccinated adults with pre-existing medical conditions. While acute SARS-CoV-2 infection is less severe among children(Chen et al., 2021; Ejaz et al., 2020; Yang et al., 2020), Multi-system Inflammatory Syndrome in Children (MIS-C)(Nakra et al., 2020; Rowley, 2020), characterized by fever, systemic inflammation, and multi-organ dysfunction following COVID-19, may occur (Consiglio et al., 2020; Noval Rivas et al., 2021). Cutaneous manifestations, such as chilblain-like lesions (CLL) or “COVID toes”, affects primarily children and young adults and are characterized by edema, erythema, violaceous discoloration on the fingers and toes, and occasional vesiculation (Baeck and Herman, 2021; Frumholtz et al., 2021; McCleskey et al., 2021). The occurrence of CLL has rapidly increased during the COVID-19 pandemic and correlates with confirmed COVID-19 incidence(McCleskey et al., 2021). Biopsy of affected tissue may confirm the diagnosis, with superficial and deep dermal lymphocytic infiltrates surrounding eccrine glands and blood vessels(Andina et al., 2020; Hernandez and Bruckner, 2020; Kanitakis et al., 2020; Massey and Jones, 2020).

CLL has been observed in patients with mild and asymptomatic COVID-19, and post-mRNA SARS-CoV-2 vaccination, but remains controversial as many patients are PCR negative without seroconversion(Kolivras et al., 2022). Chilblains may occur in the type-1 interferonopathies and affected patients with CLL can produce increased IFN-alpha, supporting that this manifestation may be associated with rapid clearance of SARS-CoV-2 infection(Hubiche et al., 2021).

Increased numbers of activated neutrophils have been described in severe COVID-19 and in MIS-C (Agbuduwe and Basu, 2020; Carter et al., 2020). Emerging evidence suggests that excessive neutrophil extracellular trap (NET) formation plays a key role in COVID-19 pathogenesis.(Barnes et al., 2020; Borges et al., 2020; Veras et al., 2020) NETs are extruded by neutrophils as a meshwork of chromatin bound to granule proteins and are synthesized during various infectious and sterile inflammatory conditions(Brinkmann et al., 2004). Infection with SARS-CoV-2 can trigger NET formation(Middleton et al., 2020). In other conditions, excessive NET formation may cause vascular injury by activating endothelial to mesenchymal transition, triggering death of endothelial cells and vascular smooth muscle cells, and promoting coagulation.(Barbosa et al., 2021; Colmenero et al., 2020; Khaddaj-Mallat et al., 2021; Leppkes et al., 2020; Zuo et al., 2021b) NET complexes have been detected in the circulation of adult patients with severe COVID-19.(Zuo et al., 2020), and widespread occlusion of small vessels by aggregated NETs has been visualized in their lung and kidneys (Radermecker et al., 2020; Schurink et al., 2020). The mechanisms by which NET remnants are elevated in circulation and tissues in COVID-19 requires further characterization. In particular, the relative contribution of enhanced NET formation versus impaired NET degradation in COVID-19 remains a matter of debate. It also remains unclear if NETs are present in asymptomatic patients infected with the virus and the natural history of elevated NET complexes after the virus is cleared and infection resolves. Furthermore, whether patients infected different variants of the virus differ in their ability to form NETs remains to be determined.

While various studies have been published in adult cohorts, the presence of NETs and the clearance mechanisms of NETs have not been systematically explored in pediatric patients displaying MIS-C or CLL. Therefore, we analyzed whether children affected with MIS-C or CLL exhibit evidence of NET dysregulation, compared with adult patients hospitalized with COVID-19, using a multi-cohort approach. Additionally, we evaluated putative mechanisms implicated in NET degradation abnormalities in subsets of COVID-19 patients. Here, we report that levels of NET remnants are elevated in MIS-C and CLL, in a geographic-dependent manner, and in association with specific disease manifestations. Pediatric and adult symptomatic COVID-19 patients displayed impaired NET degradation that did not appear to be genetically driven and was multifactorial. Furthermore, differences in NET levels were observed when comparing different strains of the virus that have been associated with different levels of disease severity and complications. These results highlight a putative pathogenic role of NETs and impairments in NET degradation in pediatric and adult subjects affected by COVID-19.

## Results

### NET remnants are elevated in circulation in children with MIS-C and CLL

We quantified NET remnants, as assessed by citrullinated histone H3 (citH3)-DNA and elastase-DNA complexes in pediatric serum/plasma samples collected in medical centers from different regions in Italy, as well as from Chile and the United States. The characteristics of these cohorts are displayed in Table 1. All patients diagnosed with CLL in Italy and the United States were negative for SARS-CoV-2 by PCR and/or negative serology and, to date, none of the patients included in the study developed evidence of chronic systemic autoimmunity post MIS-C or CLL. In the samples from Italy, levels of citH3-DNA and elastase-DNA complexes were significantly higher in children affected by CLL when compared to healthy controls (ctrls) or to MIS-C patients (Fig 1A, B). Similarly, samples obtained from children affected by CLL in the United States displayed significantly elevated levels of elastase-DNA complexes when compared to ctrls(Fig 1C). While MIS-C samples obtained from Italian cohorts did not show evidence of elevated NET remnants when compared to ctrls (Fig 1A-B), a longitudinal assessment of samples obtained from children hospitalized with MIS-C in Chile showed that citH3-DNA complexes were significantly elevated when compared to ctrls (Fig 1D).

**Figure 1.**
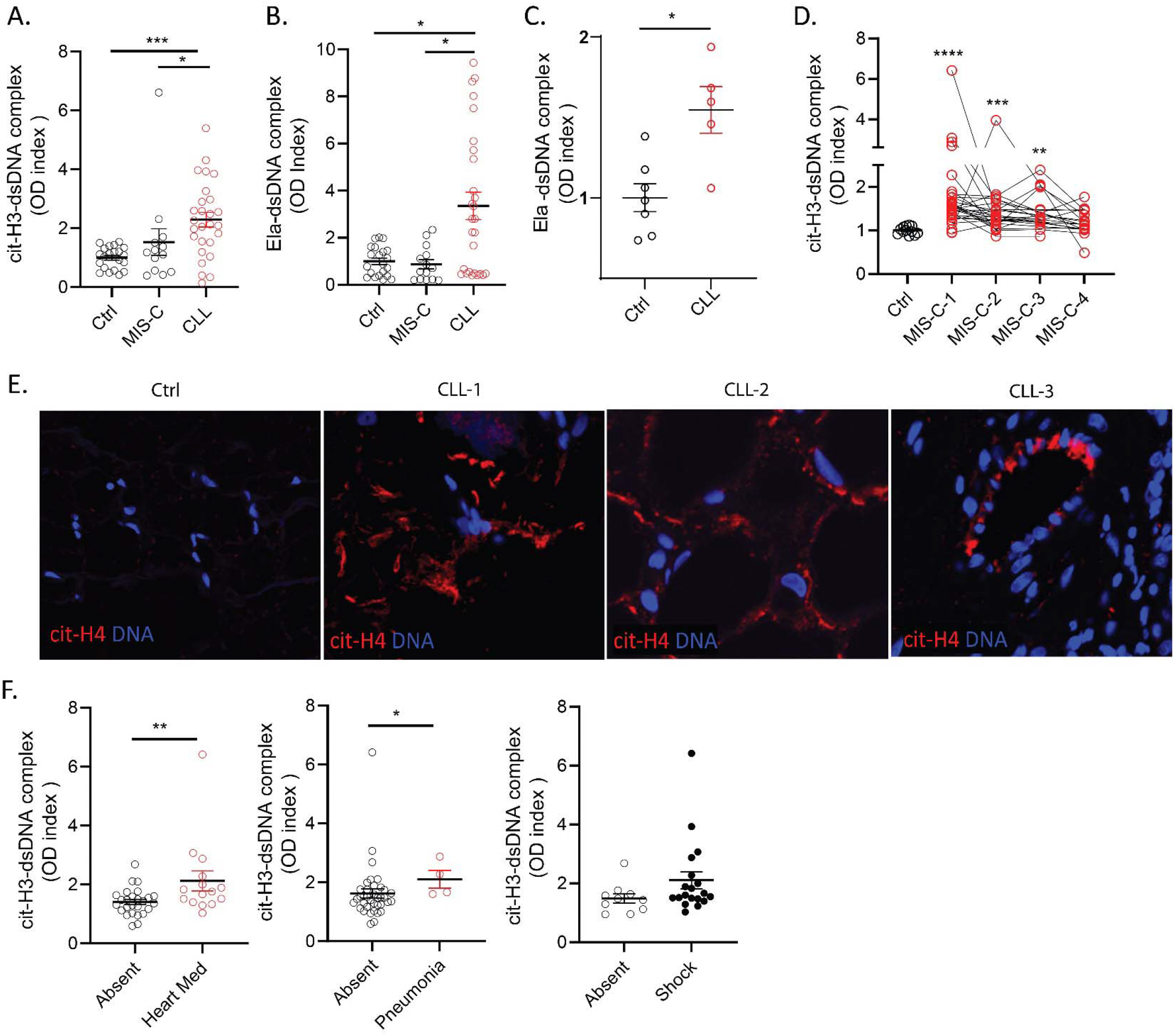
NETs are identified in MIS-C and CLL. Levels of citrullinated histone H3 (citH3)-and elastase (Ela)-DNA complexes were quantified in serum or plasma MIS-C and CLL samples obtained from (**A, B**) Italy (MIS-C n=14, CLL n= 27, ctrl n= 21), (**C**)USA (CLL n= 5, ctrl n= 5) and (**D**)Chile (MIS-C n=27, ctrl n= 12) (D). Mann-Whitney and Kruskal-Wallis analysis were performed. (**E**) Detection of citrullinated histone H4 (citH4, red) and DNA (blue) was performed in skin tissue obtained from 3 CLL patients and one ctrl (**F**) Levels of citH3-DNA complexes were correlated with the absence or present of heart medication/pressors, pneumonia or shock in Chilean MIS-C patients. Results are the mean +/- SEM. Mann-Whitney analysis were performed *p< 0.05, **p<0.01.Ctrl: controls; OD: optical density; *p<0.05, ** p<0.01, ****p<0.001.

**TABLE 1.** CHARACTERISTICS OF THE STUDY COHORTS.

|  |  |  |  |
| --- | --- | --- | --- |
| **Adult cohorts (n=280)** |  | **Chile MIS-C cohort (n=27)** |  |
| Age: 60 (19-92) |  | Age: 6 (3-11) |  |
| **Feature** | **Number (%)** | **Feature** | **Number (%)** |
| Male | 192 (68.6) | Male | 16 (59) |
| Disease severity |  | Disease severity |  |
| Critical | 168 (60) | Moderate MIS-C | 12 (44) |
| Severe | 41 (16.1) | Severe MIS-C | 15 (56) |
| Moderate | 38 (13.6) | ICU Admission | 23 (85) |
| Mild/Asymptomatic | 24 (8.6) | Pneumonia | 4 (14) |
| Site |  | Oxygen Supplementation |  |
| Brescia | 236 (84.3) | Invasive mechanical | 12 (44) |
| Monza | 25 (8.9) | Non-invasive mechanical | 0 (0) |
| ICU | 77 (27.5) | Face mask with reservoir | 0 (0) |
| Pneumonia | 230 (85) | Low flow oxygen | 2 (8) |
| Oxygen Supplementation |  | No oxygen | 13 (48) |
| Invasive mechanical | 61 (21.8) |  |  |
| Non-invasive mechanical | 57 (20.4) | **Italian MIS-C cohort (n=6)** |  |
| Face mask with reservoir | 16 (5.7) | Age: 5.8 (0.3-12) |  |
| Low flow oxygen | 40 (14.3) | **Feature** | **Number (%)** |
| No oxygen | 16 (5.7) | Male | 3 (50) |
| Unrecorded | 71 (25.4) | MIS-C | 6 (100) |
| Deceased | 45 (16.1) | Site |  |
|  |  | Turin | 3 (50) |
| **Italian CLL cohort (n=17)** |  | Naples | 3 (50) |
| Age: 13 (9-15) |  | ICU | 1 (16.7) |
| **Feature** | **Number (%)** | Pneumonia | 3 (50) |
| Male | 9 (52.9) |  |  |
| COVID toes | 17 (100) | **USA CLL cohort (n=9)** |  |
| Naples | 17 (100) | Age: 17 (14-19.5) |  |
| Hospitalization Status |  | **Feature** | **Number (%)** |
| Non-ICU | 1 (5.9) | Male | 5 (55.5) |
| Not hospitalized | 16 (94.1) | Family history of autoimmunity | 1 (11.1) |
|  |  | Recurrence(s) | 3 (33.3) |
| **Adult Omicron cohort (n=21)** |  | Systemic symptoms | 0 (0) |
| Age: 73 (64-85) |  |  | **Median (IQR)** |

| **Feature** | **Number (%)** | **Duration of skin symptoms (d)** | **145 (79)** |
| --- | --- | --- | --- |
| Omicron | 21 (100) |  |  |
| Vaccinated | 0 (0.0) |  |  |
| Male | 13 (61.9) |  |  |
| Hypertension | 6 (28.5) |  |  |
| ICU | 3 (14.2) |  |  |
| Pneumonia | 17 (80.9) |  |  |
| Critical | 16 (76.1) |  |  |

IQR: interquartile range; ICU: intensive care unit.

As levels of NETs were elevated in circulation in children presenting with CLL, we evaluated if NETs would be present in the affected tissues from biopsies obtained in the United States. All samples had histopathologic features of CLL, along with clinical correlation, and were collected after the beginning of the COVID-19 pandemic in 2020. For control skin samples, we used biopsies obtained prior to the onset of the COVID-19 pandemic. As a marker of NETs, skin specimens from CCL patients in the United States were stained for citH4, as previously described.(Carmona-Rivera et al., 2017) We detected evidence of NETs infiltrating the CLL tissues, but not the control skin (Fig 1E). CitH4-positive structures tended to localize around blood vessels and in the subcutaneous area (Fig 1E).

When assessing the associations between disease manifestations and NET remnant levels in the pediatric cohorts, levels of citH3-DNA complexes in the MIS-C cohort from Chile were significantly associated with the requirement of cardiac medications/pressors and with development of lung complications (Fig 1F). Furthermore, levels of citH3-DNA complexes displayed a trend to be elevated in those MIS-C patients that developed shock (Fig 1F). These results suggest that NETs are present in pediatric COVID-19 patients with specific disease manifestations, are associated with disease severity, and may be implicated in tissue damage in pediatric patients with CLL. The results also support geographic variation among pediatric cohorts in NET-associated disease.

### NET degradation is impaired in children with MIS-C and CLL

As NET levels were increased in pediatric patients with CLL and MIS-C, we tested whether NET degradation mechanisms were impaired in these populations. Ctrl neutrophils were stimulated with phorbol myristate acetate (PMA) for 4 h to generate NETs, followed by addition of 5 % serum or plasma samples from ctrls, MISC or CLL patients for 16 h to identify NET degraders versus non-degraders. Italian MIS-C and CLL samples displayed a bimodal distribution, where 60 % of MIS-C and 17% of CLL samples did not significantly degrade the NETs (Fig 2A-C). In contrast, 97% of MIS-C samples from Chile and all the pediatric CLL samples from the United States displayed significantly impaired NET degradation when compared to ctrl samples (Fig 2A-C). Indeed, the degree of NET degradation differed between regions. MIS-C samples from Chile displayed significant impairments in ability to degrade NETs when compared to Italian samples (Figure 2A). Furthermore, the ability to degrade NETs by serum samples from CLL patients from the United States was significantly impaired when compared to those CLL samples obtained from Italy (Figure 2B). These results support that MIS-C and CLL patients display impairments in their ability to degrade NETs in vitro in a geographic-dependent manner.

**Figure 2.**
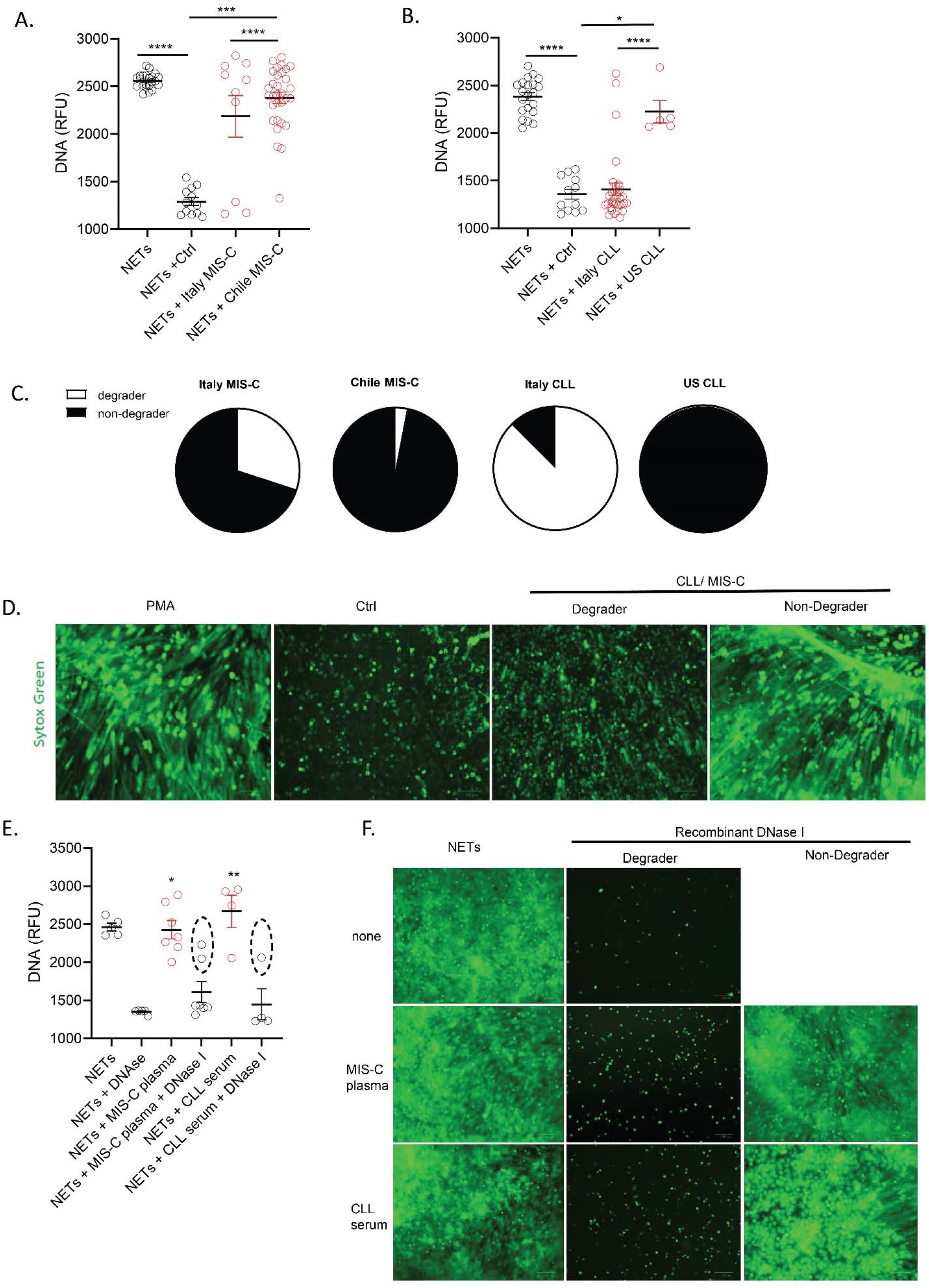
Impaired NET degradation in MIS-C and CLL samples. NET degradation capabilities were measured in serum or plasma from MIS-C samples obtained from **(A)** Italy (MIS-C n=12, control n= 12) and Chile (MIS-C n=27) and CLL obtained from (**B**) Italy (CLL n= 27, control n= 12) and US (n=5). Results are the mean +/- SEM. Kruskal-Wallis analysis was performed. (**C**) Pie charts representing the proportion of degrader (white) and non-degrader (black) per cohort. (**D**) Representative images of PMA-generated NETs incubated with serum or plasma from control or MIS-C, CLL patients. DNA is detected by Sytox green and scale bar is 100um. (**E**) NET degradation capabilities were measured in serum or plasma of MIS-C (n=7) and CLL (n= 4) samples in the presence or absence of recombinant DNase 1. Samples within dashed circle are those that did not respond to treatment with DNase1. Kruskal-Wallis analysis was performed. (**F**) Representative images of PMA generated NETs incubated with serum or plasma from MIS-C, CLL patients in the presence or absence of recombinant DNase 1. DNA is detected by Sytox green and scale bar is 100um; *p< 0.05, **p<0.01, ***p<0.001, ****p<0.0001

To elucidate the possible mechanisms involved in NET degradation impairment of MIS-C and CLL samples, we added exogenous recombinant DNase1 to the sera. We hypothesized that if the mechanism of impaired NET degradation involved improper function of the endogenous nucleases, adding exogenous nucleases would lead to restoration of the clearance mechanisms. Indeed, addition of exogenous DNase1 to MIS-C or CLL samples resulted in a significant increase in NET degradation in most samples when compared to MIS-C or CLL samples in the absence of exogenous DNase1 (Fig 2). However, 29% of the MIS-C samples and 25% of the CLL samples did not show evidence of restored clearance capacity with exogenous nucleases (Fig 2). These results suggest that some patients may have circulating DNase1 inhibitors or other mechanisms that enhance inability of nucleases to effectively access the NETs for degradation.

### Impairments in NET degradation in MIS-C and CLL are multifactorial

Impaired NET degradation can be secondary to a variety of genetic and acquired factors. To test whether genetic factors that may alter the function of nucleases are involved in the impaired NET degradation observed in pediatric COVID-19, we performed whole genome association analysis (WGS) within a subset of subjects and conducted variant association analysis for genes encoding for enzymes or molecules involved in DNA degradation (*DNASE1, DNASE1L3, DNASE1L1, DNASE1L2, DNASE2B, SERPINB1, TREX1* and *TREX2)*. The analysis did not reveal any candidate homozygous rare or low-frequency (MAF< 0.1) variants associated with the non-degrader when compared to the degrader group (Supplementary Table 1). Furthermore, we did not observe heterozygous common (MAF >0.1) variants with distribution attributed to the degrader or non-degrader group (Supplementary Table 1). Overall, a targeted genetic association analysis did not yield evidence associations between genetic alterations in nucleases and impaired NET degradation in the subjects. We therefore investigated other factors that may be involved in aberrant NET degradation.

Complement activation and its deposition in NETs has been linked to impairment in NET degradation.(Leffler et al., 2012; Skendros et al., 2020) PMA-induced healthy ctrl NETs incubated with serum or plasma from MIS-C subjects, but not from CLL or ctrl subjects, stained positive for C1q (Fig 3A), suggesting that complement activation may play a role in impairing NET degradation in MIS-C. G-actin is a natural inhibitor of DNase1 that has also been reported to impair NET degradation(Hakkim et al., 2010). Levels of G-actin in the samples from the Chilean MIS-C cohort were significantly elevated when compared to ctrls (Fig 3B), while they were not elevated in patients with CLL (Fig 3B). Anti-NET autoantibodies (autoAbs) have been previously associated with impairments in NET degradation in conditions such as systemic lupus erythematosus, and these autoAbs have been reported in adult COVID-19 patients(Hakkim et al., 2010; Zuo et al., 2021a). Therefore, we tested for their presence in MIS-C or CLL samples. We detected IgG binding to NETs in CLL but not in MIS-C samples (Fig 3C), in association with impaired degradation (r= 0.6012, p= 0.0006; Fig 3D). Overall, different factors that include complement activation, increased level of G-actin, and the development of anti-NET Abs appear to contribute to impaired NET degradation in pediatric cases of COVID-19.

**Figure 3.**
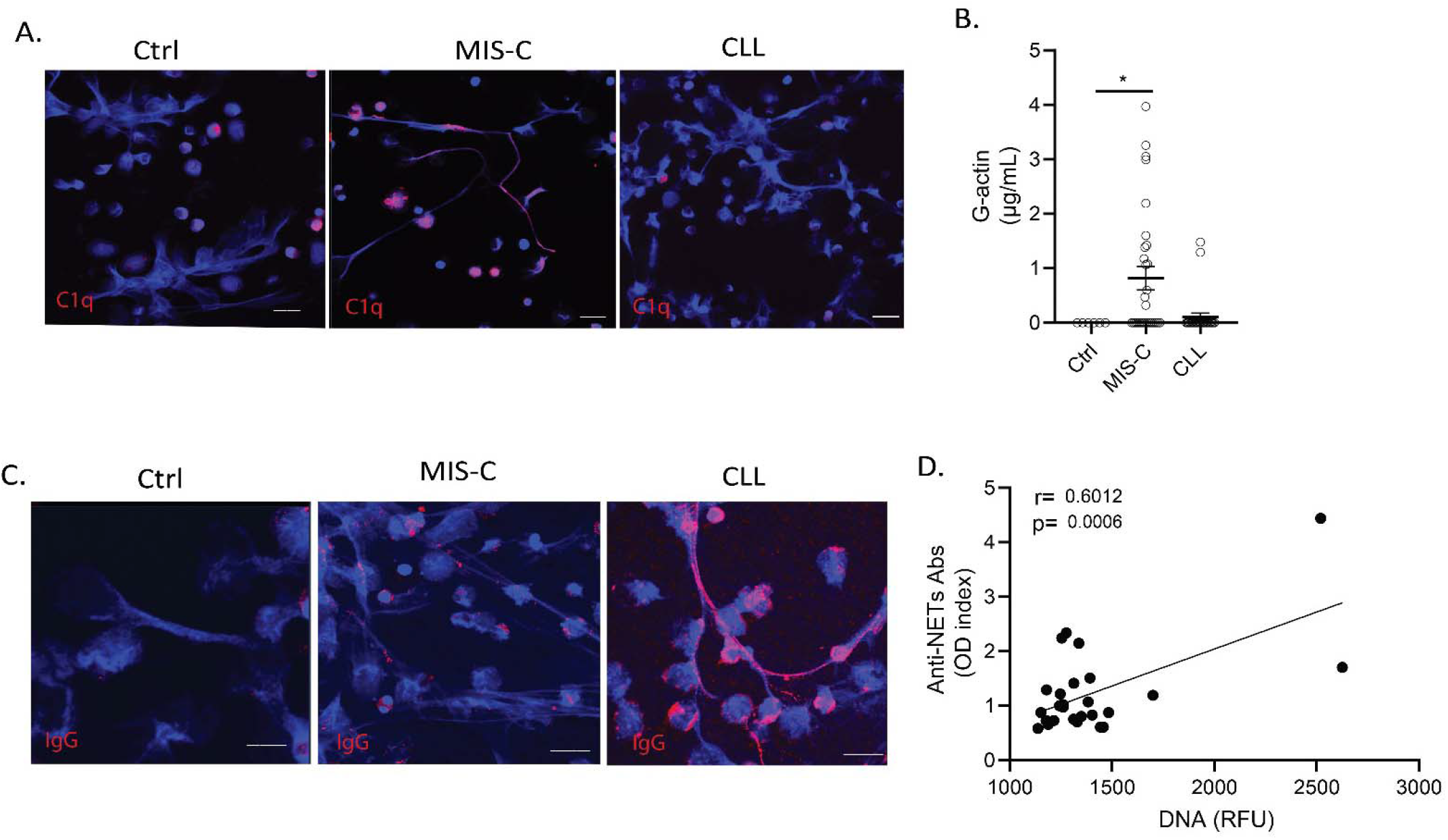
Multiple factors impair NET degradation in MIS-C and CLL. **(A)** Confocal images of immunofluorescence analysis of C1q (red) in PMA-generated NETs after incubation with serum or plasma from MIS-C or CLL patients. DNA is detected in blue and scale bar is 10um. (**B**) Levels of circulating G-actin measured in MIS-C (n=27) and CLL (n=26) samples. Results are the mean +/- SEM. Kruskal-Wallis analysis was performed *p< 0.05. (**C)** Confocal images of immunofluorescence analysis of immunoglobulin G (IgG) (red) in PMA generated NETs after incubation with serum or plasma from MIS-C or CLL patients. Scale bar is 10um. (**D**) Correlation analysis of levels of anti-NET antibodies and degradation capabilities, Pearson analysis was used, n=27.

### Persistently elevated NET remnants are detected in adult patients with symptomatic COVID-19 in association with disease manifestations

To compare the results obtained in the pediatric population with adults that developed COVID-19, we validated the findings and quantified NET remnants (citH3-DNA and elastase-DNA complexes) in plasma samples obtained from adult patients admitted to medical centers in different cities in Italy (Brescia, Turin and Monza; Table 1). Complexes of citH3-DNA and elastase-DNA were significantly elevated in these patients (Fig 4A-B). NET complexes were measured in serum samples collected at different days following the day of hospitalization. A significant increase in cit-H3-DNA complexes was detected in samples collected up to 25 days from the day of admission to the hospital (Fig 4 C-D). Since infection with SARS-CoV-2 can lead to symptomatic and asymptomatic disease, we asked whether NET complexes would be present in asymptomatic patients infected with SARS-CoV-2. The analysis indicated that NET remnants were significantly lower in asymptomatic subjects, compared to symptomatic patients (Fig 4E). Many COVID-19 patients continue to have clinical complications months after initial diagnosis(Bagnato et al., 2020; Higgins et al., 2021; Huang et al., 2021a). We found that circulating complexes of citH3-DNA were significantly elevated even 3 months after infection diagnosis, when compared to ctrls (Fig 4F). Indeed, there was no significant difference between initial and 3-month post diagnosis levels of citH3-DNA complexes, although some patients displayed reductions (Fig 4F). These results suggest that increased NET levels in adults with COVID-19 are associated with the presence of symptoms and can persist for several months post-initial infection.

**Figure 4.**
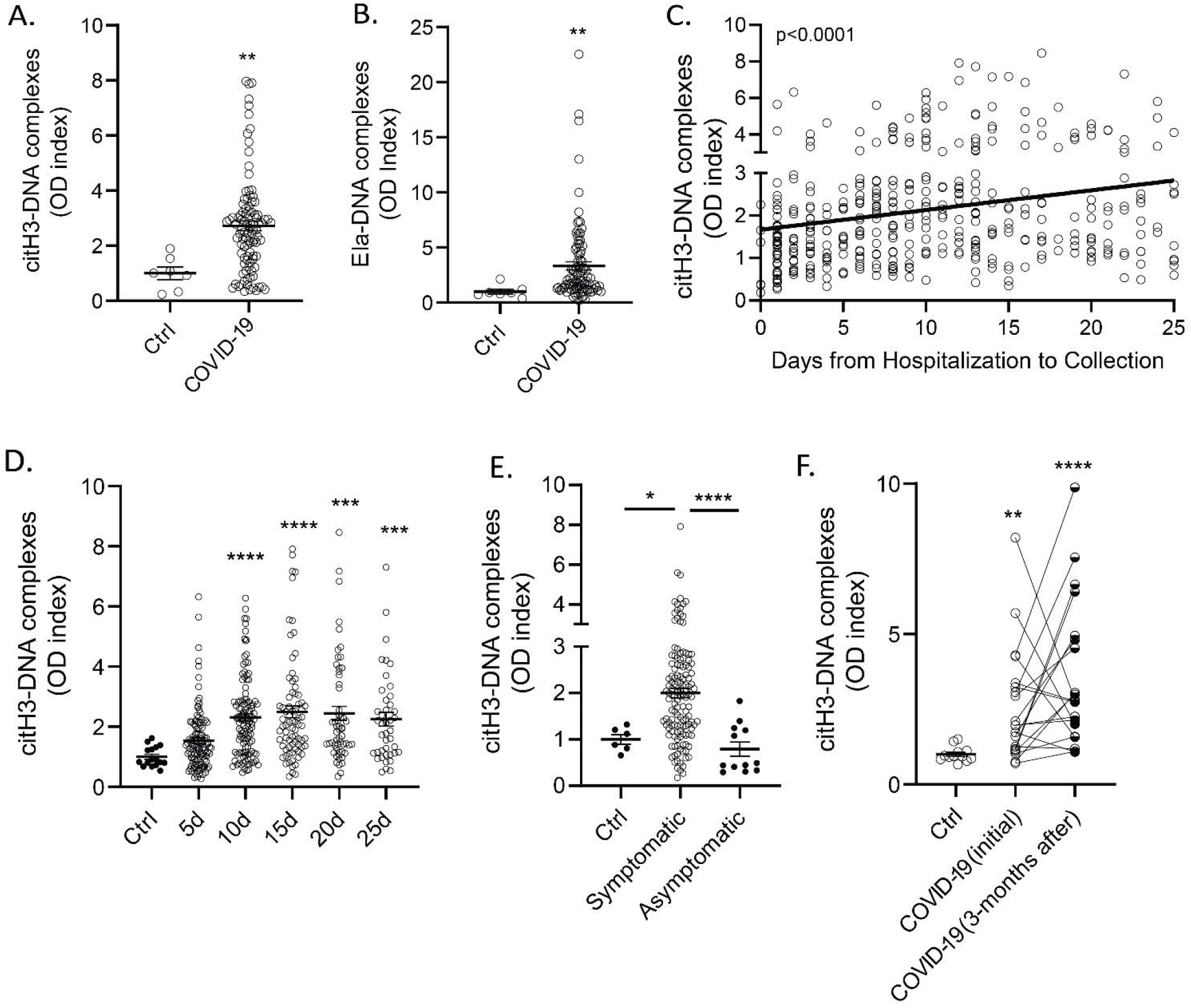
NET remnants are present in plasma and serum of COVID-19 patients from Italy. Plasma levels of (**A**) citrullinated histone H3 (citH3) and (**B**) elastase (Ela)-DNA complexes were measured in COVID-19 samples obtained from Brescia, Italy (n= 99, control n= 7), Mann-Whitney was used. (**C, D**) Serum levels of citrullinated histone H3-DNA complexes were elevated in samples obtained up to 25 days (5d n= 118, 10d n= 117, 15d n= 77, 20d n=57, 25d n= 42) of hospitalization and sample collection. Kruskal-Wallis analysis was used. Levels of citH3-DNA complexes were measured in (**E**) symptomatic (n= 77) and asymptomatic COVID-19 (n= 12) samples. (**F**) Levels of citH3-DNA complexes at initial (n=20) and 3 months after diagnosis of infection with SARS-CoV-2. Results are the mean +/- SEM. Kruskal-Wallis analysis was used, *p< 0.05, **p<0.01, *** p<0.001, ****p<0.0001. OD: optical density; ctrl:control; d:days from diagnosis.

### NETs associate with disease severity in adult COVID-19

To gain further insight into the possible role of NETs in the symptoms and complications of COVID-19, we analyzed multiple parameters that were specifically available for the adult COVID-19 Brescia cohort. We found that serum levels of citH3-DNA complexes were significantly elevated in patients with COVID-19 that were in severe or critical condition (Fig 5A), those who required intensive care unit (ICU) admission (Fig 5B), those with pneumonia (Fig 5C), and those who required high flow oxygen when compared to those with no need for oxygen supplementation (Table 2). Serum levels of citH3-DNA were not associated with sex, age, death, diabetes, cardiovascular disease, chronic heart failure, and other comorbidities (Table 2). Of note, serum levels of citH3-DNA were significantly decreased in patients who developed COVID-19 and had underlying chronic hypertension, chronic kidney disease or history of solid malignancy (Table 2). In the adult patient population with COVID-19, we found that levels of serum citH3-DNA most consistently associated with disease severity, while levels of elastase-DNA followed less consistent and sometimes opposite trends compared to the citH3-DNA levels (Supplementary Table 2).

**Figure 5.**
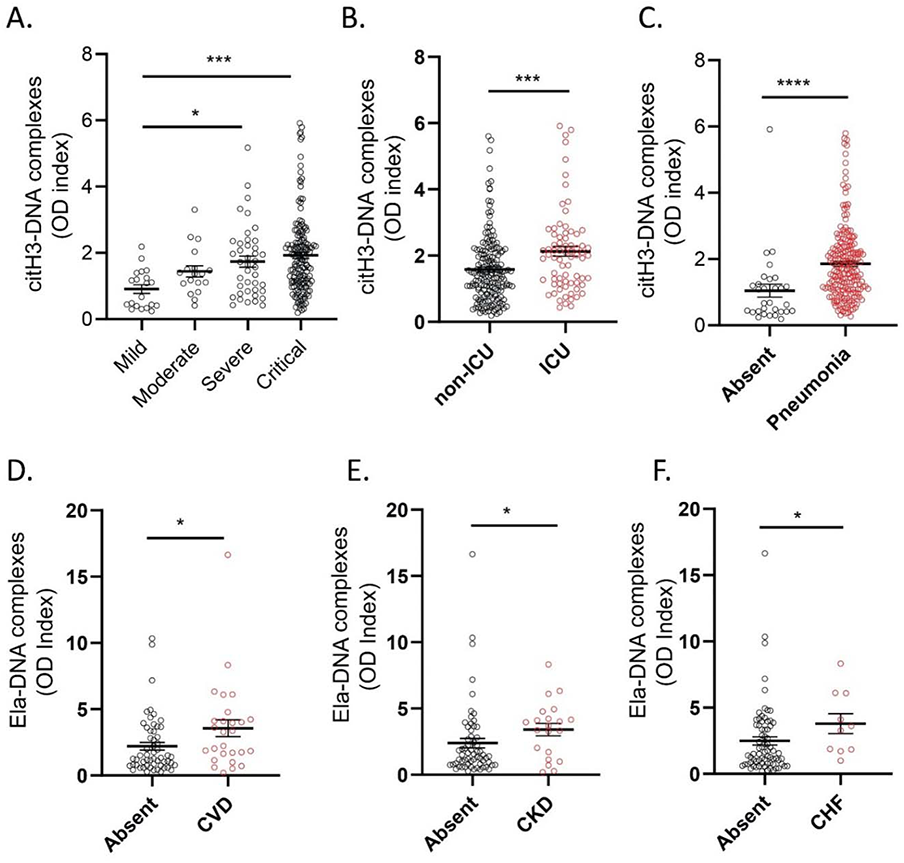
NET remnants are present in plasma and serum of adult COVID-19 patients and correlate with severity and comorbidities. Serum levels of (**A**) citrullinated histone H3 (citH3) were elevated in critical patients (mild n= 20, moderate n=18, severe n= 41, critical n=156). Kruskal-Wallis analysis was used. (**B**) COVID-19 patients in intensive care unit (ICU) displayed elevated levels of citH3-DNA complexes (non-ICU n= 175, ICU n= 73) (**C**) Patient with pneumonia displayed elevated levels of citH3-DNA complexes (absent n= 30, pneumonia n= 203), Mann-Whitney was used. (**D**) Plasma levels of elastase-DNA complexes (Ela-DNA) were elevated COVID-19 samples with comorbidities such as cardiovascular disease (CVD) (absent n= 56, CVD n= 27), (**E**) chronic kidney disease (CKD) (absent n= 62, CKD n= 21), and **(F**)congestive heart failure (CHF) (absent n= 73, CHF n= 10). Results are the mean +/- SEM. Mann-Whitney was used. *p<0.05, ***p<0.001, ****p<0.0001. OD: optical density.

**TABLE 2.** CORRELATIONS BETWEEN CITRULLINATED HISTONE H3: DNA COMPLEXES AND CLINICAL VARIABLES IN SERUM SAMPLES.

| **FEATURE** | **VARIABLE (N);**<br>**MEDIAN** | **VARIABLE (N);**<br>**MEDIAN** | **P Value** |
| --- | --- | --- | --- |
| Sex | F (71); 1.469 | M (176); 1.536 | 0.56224 |
| Age | <60 (120); 1.531 | >60 (127); 1.487 | 0.81405 |
| Severity | Mild (20); 0.876 | Moderate (18);<br>1.366 | 0.02047 |
| Severity | Mild (20); 0.876 | Severe (41); 1.642 | 0.00087 |
| Severity | Mild (20); 0.876 | Critical (156); 1.721 | 0.00003 |
| Oxygen needs | None (14); 1.383 | Low flow (83); 1.545 | 0.45673 |
| Oxygen needs | None (14); 1.383 | High flow (22); 2.154 | 0.01889 |
| Oxygen needs | None (14); 1.383 | Intubation (31);<br>2.227 | 0.06737 |
| Oxygen needs | None (14); 1.383 | Tracheostomy (27);<br>2.016 | 0.22420 |
| Pneumonia | Absent (30); 0.679 | Present (203); 1.642 | 1.62E-06 |
| Renal insufficiency | Absent (227); 1.487 | Present (6); 1.764 | 0.48807 |
| Death | Alive (200); 1.474 | Deceased (43);<br>1.609 | 0.96948 |
| Need for ICU | ICU (73); 2.016 | non-ICU (174); 1.402 | 0.00040 |
| Comorbidities | Absent (183); 1.504 | Present (45); 1.722 | 0.32151 |
| CVD | Absent (178); 1.626 | Present (50); 1.305 | 0.27970 |
| Hypertension | Absent (137); 1.643 | Present (91); 1.480 | 0.03985 |
| Diabetes mellitus | Absent (185); 1.556 | Present (43); 1.504 | 0.48271 |
| CHF | Absent (209); 1.584 | Present (19); 1.168 | 0.16524 |
| CKD | Absent (186); 1.716 | Present (42); 0.995 | 0.00006 |
| Obesity | Absent (207); 1.527 | Present (21); 1.926 | 0.11416 |
| Hematologic malignancy | Absent (177); 1.654 | Present (51); 1.285 | 0.07943 |
| Solid malignancy | Absent (204); 1.630 | Present (21); 1.197 | 0.03401 |
| Neurologic/Psychiatric disease | Absent (182); 1.560 | Present (46); 1.492 | 0.29857 |
| Autoimmunity | Absent (180); 1.643 | Present (48); 1.194 | 0.05443 |
| Chronic Respiratory disease | Absent (206); 1.531 | Present (22); 1.629 | 0.85031 |
| Liver disease | Absent (210); 1.551 | Present (18); 1.425 | 0.61260 |
| Solid organ transplant | Absent (211); 1.584 | Present (17); 0.882 | 0.01140 |

ICU: intensive care unit; CVD: cardiovascular disease; CHF: congestive heart failure; CKD: chronic kidney disease

Plasma levels of elastase-DNA and citH3-DNA were not associated with sex, age, death, obesity, autoimmune disorder, or neurological-psychiatric disease (Table 3, Supplementary Table 3). Both plasma elastase-DNA and citH3-DNA levels were associated with having an underlying diagnosis of cardiovascular disease (Fig 5D), chronic kidney disease (Fig 5E), or history of solid organ transplantation (Table 3 and Supplementary Table 3). Plasma elastase-DNA was also associated with congestive heart failure (Fig 5F), while plasma citH3-DNA was associated with liver disease (Supplementary Table 3). Of note, levels of NET remnants were similar in adult and pediatric COVID-19 cohorts. Overall, levels of NET remnants were associated with specific disease manifestations and disease severity in adult subjects diagnosed with COVID-19.

**TABLE 3.** CORRELATIONS BETWEEN ELASTASE: DNA COMPLEXES AND CLINICAL VARIABLES IN PLASMA SAMPLES.

| FEATURE | VARIABLE (N);<br>MEDIAN | VARIABLE (N);<br>MEDIAN | P Value |
| --- | --- | --- | --- |
| FEATURE | MEDIAN |  | P Value |
| Sex | F (17); 2.029 | M (74); 1.669 | 0.51133 |
| Age | <60 (36); 2.554 | >60 (55); 1.701 | 0.39184 |
| Severity | Mild (2); 2.516 | Moderate (3); 3.854 | 0.80000 |
| Severity | Mild (2); 2.516 | Severe (14); 3.786 | 0.26667 |
| Severity | Mild (2); 2.516 | Critical (66); 1.452 | 0.33611 |
| Oxygen needs | None (1); 2.804 | Low flow (22); 2.046 | 0.86957 |
| Oxygen needs | None (1); 2.804 | High flow (5); 1.740 | 1.00000 |
| Oxygen needs | None (1); 2.804 | Intubation (14);<br>1.157 | 0.66667 |
| Oxygen needs | None (1); 2.804 | Tracheostomy (21);<br>1.155 | 0.36364 |
| Pneumonia | Absent (7); 3.324 | Present (77); 1.551 | 0.05829 |
| Renal Insufficiency | Absent (81); 1.701 | Present (3); 3.280 | 0.42637 |
| Death | Alive (74); 1.744 | Deceased (13);<br>1.699 | 0.75238 |
| Need for ICU | ICU (41); 1.155 | non-ICU (49); 2.804 | 0.00842 |
| Comorbidities | Absent (77); 1.872 | Present (6); 0.574 | 0.00532 |
| CVD | Absent (56); 1.296 | Present (27); 3.280 | 0.01890 |
| Hypertension | Absent (44); 1.363 | Present (39); 2.029 | 0.12921 |
| Diabetes Mellitus | Absent (62); 1.721 | Present (21); 1.841 | 0.92905 |
| CHF | Absent (73); 1.509 | Present (10); 3.444 | 0.03649 |
| CKD | Absent (62); 1.452 | Present (21); 3.688 | 0.01622 |
| Obesity | Absent (74); 1.721 | Present (9); 2.888 | 0.37558 |
| Hematologic malignancy | Absent (55); 1.939 | Present (28); 1.452 | 0.11756 |
| Solid malignancy | Absent (73); 1.841 | Present (10); 1.326 | 0.38194 |
| Neurologic/Psychiatric Disease | Absent (62); 1.594 | Present (21); 2.029 | 0.45389 |
| Autoimmunity | Absent (58); 1.530 | Present (25); 2.428 | 0.46567 |
| Chronic respiratory disease | Absent (74); 1.744 | Present (9); 1.640 | 0.99416 |
| Liver disease | Absent (75); 1.551 | Present (8); 2.576 | 0.44499 |
| Solid organ transplant | Absent (73); 1.509 | Present (10); 3.522 | 0.02858 |

ICU: intensive care unit; CVD: cardiovascular disease; CHF: congestive heart failure; CKD: chronic kidney disease

|  |  |  |  |
| --- | --- | --- | --- |
| Sex | F (44); 1711 | M (86); 1754 | 0.95880 |
| Age | <60 (72); 1775 | >60 (58); 1707.5 | 0.37985 |
| Severity | Mild (17); 1454 | Moderate (8); 1887.5 | 0.00901 |
| Severity | Mild (17); 1454 | Severe (13); 2542 | 0.00284 |
| Severity | Mild (17); 1454 | Critical (89); 1714 | 0.12018 |
| Oxygen needs | None (8); 2269.5 | Low flow (43); 1854 | 0.23860 |
| Oxygen needs | None (8); 2269.5 | High flow (15); 1792 | 0.19011 |
| Oxygen needs | None (8); 2269.5 | Intubation (20); 1660.5 | 0.02131 |
| Oxygen needs | None (8); 2269.5 | Tracheostomy (20); 1707.5 | 0.07916 |
| Pneumonia | Absent (17); 1454 | Present (109); 1792 | 0.03938 |
| Renal Insufficiency | Absent (121); 1714 | Present (5); 2147 | 0.36491 |
| Death | Alive (107); 1775 | Deceased (23); 1532 | 0.09519 |
| Need for ICU | ICU (48); 1696 | non-ICU (82); 1787.5 | 0.12800 |
| Comorbidities | Absent (87); 1792 | Present (40); 1623.5 | 0.26334 |
| CVD | Absent (112); 1754 | Present (15); 1714 | 0.94342 |
| Hypertension | Absent (91); 1691 | Present (36); 1828.5 | 0.32103 |
| Diabetes Mellitus | Absent (107); 1739 | Present (20); 1758 | 0.87379 |
| CHF | Absent (121); 1739 | Present (6); 2192.5 | 0.29581 |
| CKD | Absent (123); 1739 | Present (4); 2352.5 | 0.08953 |
| Obesity | Absent (114); 1754 | Present (13); 1593 | 0.72936 |
| Hematologic malignancy | Absent (110); 1689.5 | Present (17); 2017 | 0.01501 |
| Solid malignancy | Absent (117); 1691 | Present (10); 2279.5 | 0.02523 |
| Neurologic/Psychiatric disease | Absent (110); 1689.5 | Present (17); 2583 | 0.01091 |
| Autoimmunity | Absent (117); 1767 | Present (10); 1682.5 | 0.74049 |
| Chronic Respiratory Disease | Absent (115); 1767 | Present (12); 1579 | 0.27479 |
| Liver Disease | Absent (121); 1739 | Present (6); 1930 | 0.68664 |
| Solid Organ Transplant | Absent (125); 1741 | Present (2); 1736.5 | 1.00000 |

### NET degradation is impaired in adult patients with COVID-19

We analyzed NET degradation capability of serum samples obtained from Italian adult COVID-19 patients. Similar to the pediatric population, impairments in NET degradation were significantly increased in the serum samples from adult COVID-19 patients when compared to ctrls (Fig 6A), and the impairment was significantly higher in symptomatic COVID-19 patients when compared to those who were asymptomatic at diagnosis (Fig 6A). In adults, 54% of the COVID-19 samples tested displayed impairments in NET degradation (Fig 6A-B). We investigated whether the lack of NET degradation persisted after acute infection period had passed. To address this, we tested the NET degradation capacity of serum samples from 20 COVID-19 patients at initial diagnosis and 3 months later. A significant improvement in NET degradation was observed 3 months after initial infection with SARS-CoV-2, with a reduction of non-degraders from 65 to 25% of the samples tested (Fig 6C).

**Figure 6.**
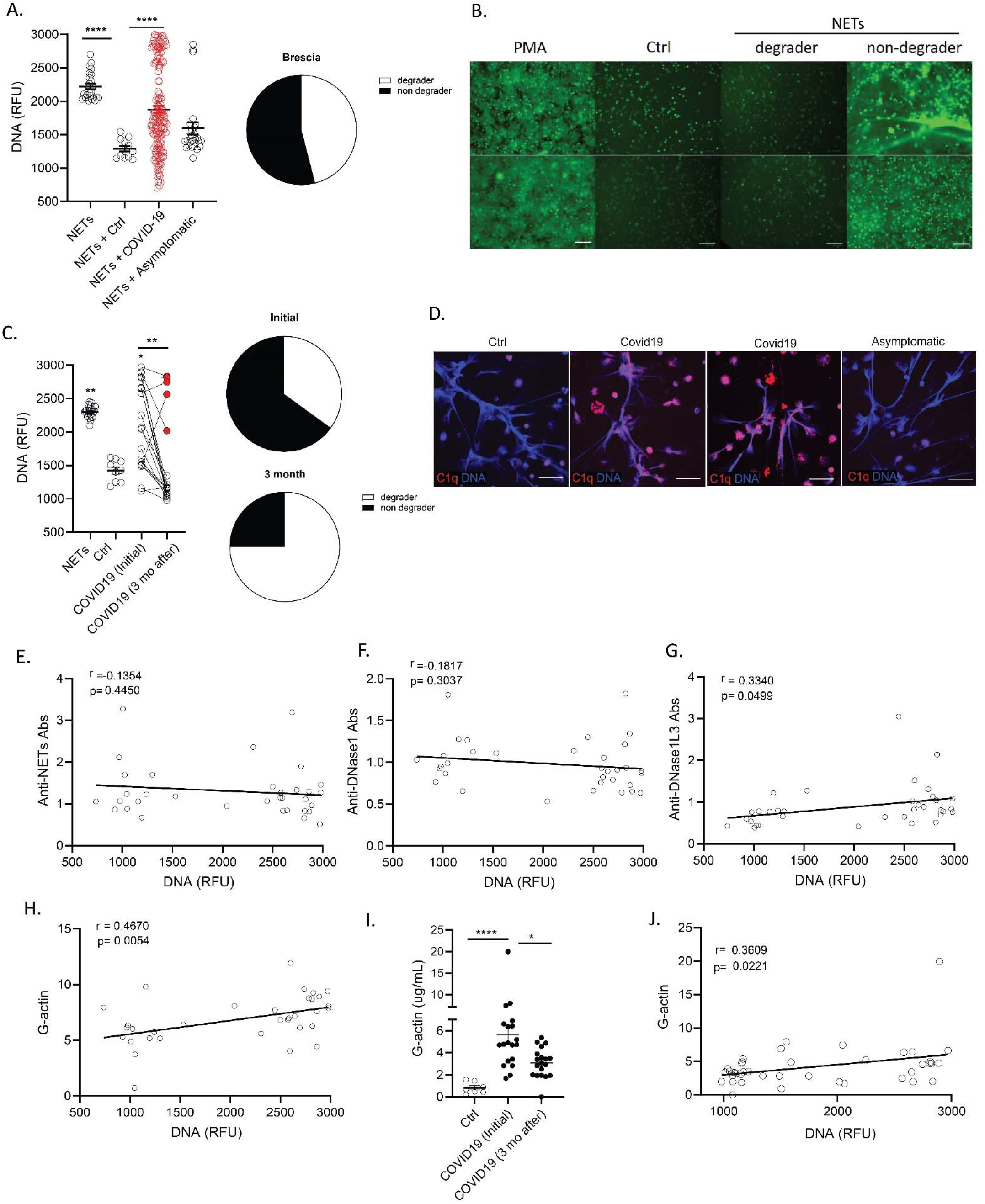
G-actin associates to decreased degradation of NETs in adult Italian COVID-19 subjects. **(A)** NET degradation capabilities were measured in COVID-19 serum samples obtained from Brescia Italy (COVID-19 n=153, asymptomatic n= 26, control n=12), pie chart depicting the proportion of degrader (white) and non-degrader (black). Results are the mean +/- SEM. Kruskal-Wallis analysis was performed. (**B**) Representative images of PMA generated NETs incubated with serum from control or COVID-19 patients. DNA is detected by Sytox green and scale bar is 100um. (**C**) NET degradation capabilities were assessed in twenty patients at initial infection and 3 months (3 mo) after. Pie charts depicting the proportion of degrader (white) and non-degrader (black) at initial and 3 months after infection with SARS-CoV-2. Kruskal-Wallis analysis was performed. (**D**) Confocal images of immunofluorescence analysis of C1q deposition (red) in PMA-generated NETs after incubation with serum from symptomatic and asymptomatic COVID-19 patients. DNA is detected in blue and scale bar is 10um. Pearson correlation analysis of (**E**) levels of anti-NET, (**F**) anti-DNase1, (**G**) anti-DNase1L3 Abs and (**H**) G-actin measured in serum from COVID-19 patients with NET degradation capabilities (DNA). (**I**) Levels of serum G-actin in COVID-19 patients (n=20) at initial and 3 months after diagnosis of infection with SARS-CoV-2. Kruskal-Wallis analysis was performed. (**J**) Pearson correlation analysis of G-actin measured in serum from COVID-19 patients at initial and 3 months after infection with SARS-CoV-2 with NET degradation capabilities (DNA). *p< 0.05, **p<0.01, ****p<0.0001; ctrl:contrl; mo:months; RFU: relative fluorescence units.

WGS analysis was also performed in adult patients to determine if there were putative genetic contributions of rare or common variants within genes associated with DNA degradation and their link to NET degradation impairments in adult COVID-19 patients from Italy. No common (minor allele frequency (MAF) >0.1) variants were detected with distribution toward degraders or non-degraders. However, a few candidates of loss of function (LOF) homozygous rare (MAF<0.1) variants and a very rare heterozygous frameshift variant in *DNASE2* were detected in non-degraders (Supplementary Table 4).

As we found evidence of C1q deposition in NETs exposed to MIS-C serum (but not CLL serum), we tested if C1q deposition in NETs occurred in adult patients with COVID-19. Confocal microscopy analysis demonstrated C1q deposition in NETs after incubation with symptomatic adult COVID-19 sera but not in NETs incubated with healthy control or asymptomatic adult COVID-19 samples (Fig 6D). Anti-NET autoAbs have been previously reported in adults with COVID-19(Zuo et al., 2021a) and we found them to associate with lack of NET degradation in pediatric CLL samples (Fig 2D). However, we did not find significant levels of anti-NETs autoAbs and the levels of anti-NET autoAbs measured in adult COVID-19 samples from Italy did not correlate with impairments in NET degradation (Fig 6E). We quantified autoAbs against the most abundant nucleases in serum, DNase1 and DNase1L3. Levels of autoAbs against DNase1L3, but not against DNase, were associated with impairments in NET degradation (Fig 6 F-G), suggesting that anti-DNase1L3 Abs may be implicated in impairing clearance of NETs in adult patients with COVID-19. We also measured the levels of G-actin and found a significant correlation between levels of G-actin and impaired NET degradation in adult COVID-19 serum samples (Fig 6 H). Furthermore, longitudinal analysis of COVID-19 patients demonstrated that the levels of G-actin were significantly reduced after 3 months from the initial infection diagnosis (Fig 6I) and these levels significantly correlated (r= 0.3609, p= 0.0221) with impairments in NET degradation (Fig 6J). Similar to the pediatric samples, these results indicate that the significant impairments in NET degradation in adults with COVID-19 are multifactorial and may be associated with the presence of autoAbs against DNase1L3 and elevated levels of G-actin present in COVID-19 serum.

When assessing associations with COVID-19 manifestations and impaired NET degradation in adults, we found associations with moderate and severe clinical phenotype compared to mild phenotype (Fig 7A, Table 4). In addition, NET degradation was significantly impaired in patients with pneumonia, previous neurologic manifestations, and history of cancer (Fig 7 B-D).

**Figure 7.**
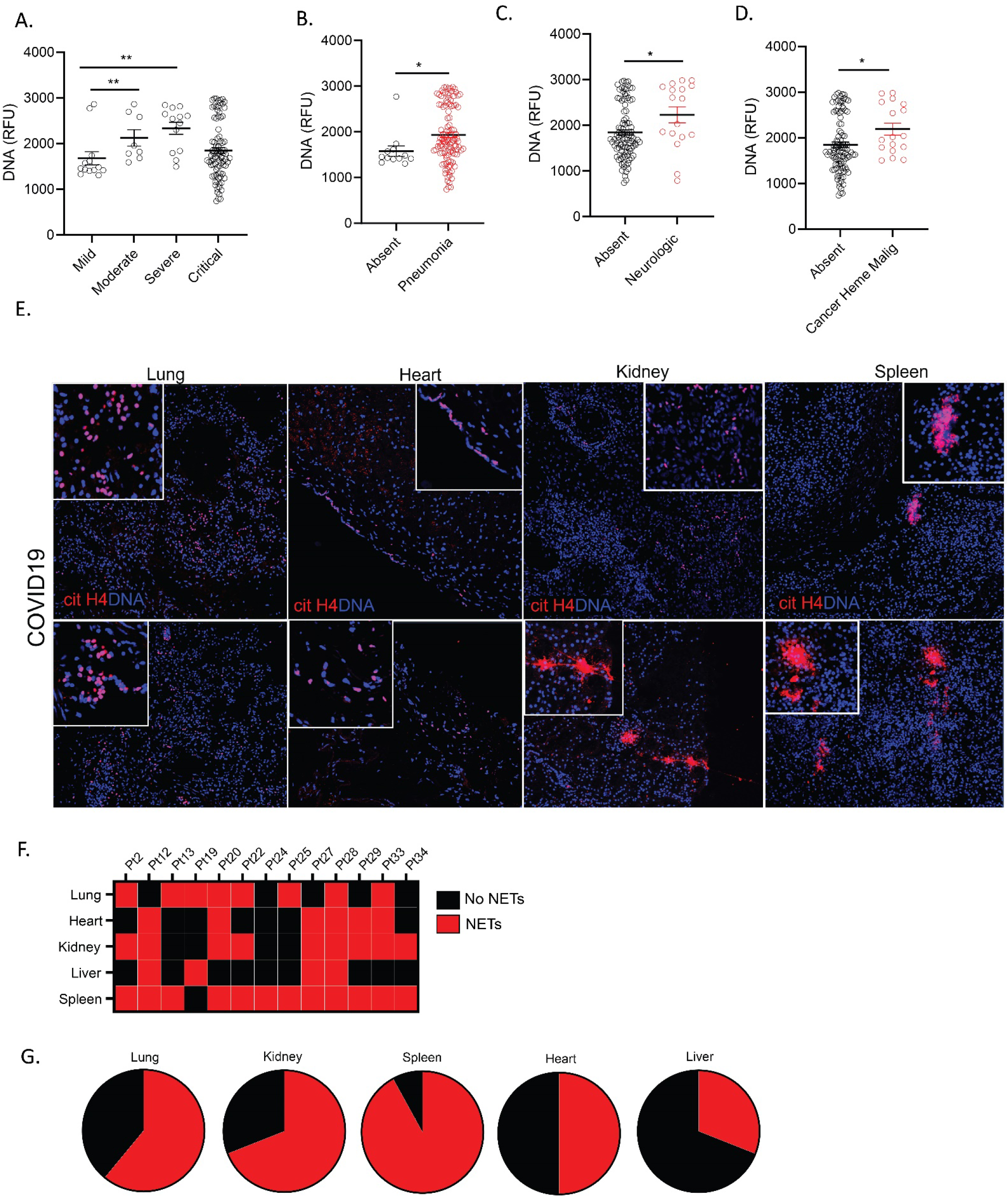
Impairment in NET degradation correlates with comorbidities, while NETs are detected in pulmonary and extrapulmonary tissues in COVID-19. (**A**) Decreased serum NET degradation capabilities in severe COVID-19 patients (mild n= 17, moderate n= 8, severe n= 13, critical n=89). Kruskal-Wallis analysis was used. COVID-19 patients with (**B**) pneumonia (absent n=17, pneumonia n=109), (**C**) neurologic manifestations (absent n=103, neurologic n=17), and (**D**) malignancy (absent n=110, malignancy n=17) displayed decreased capabilities of NET degradation. Results are the mean +/- SEM. Mann-Whitney was used. (**E**) Representative confocal images of citrullinated histone H4 (citH4, red) and DNA (blue) detected in lung, heart, kidney, and spleen tissues obtained from post-mortem COVID-19 patient. (**F**) Summary of tissue NET detection in each patient (n=13). (**G**) Pie charts depicting global NET detection per tissue analyzed. Red indicates positive to citH4 (NETs), black indicates no presence of citH4 signal (no NETs); *p<0.05, ***p<0.001.

**TABLE 4.** CORRELATIONS BETWEEN IMPAIRED NET DEGRADATION AND CLINICAL VARIABLES. ICU: intensive care unit; CVD: cardiovascular disease; CHF: congestive heart failure; CKD: chronic kidney disease

### NETs are present in COVID-19 patient tissues

Since NETs appear to be associated with various manifestations of COVID-19 and correlate with disease severity, we analyzed if we could detect them in various tissues (lung, heart, spleen, kidney and liver) available at autopsy from 13 patients who died from complications of this disease. We used cit-H4 marker as a readout of tissue NETs, as described above for the CLL samples. We detected NETs in all these tissues, with variability among patients, including one subject who had detectable NETs in all tissues tested (Fig 7E). The organs that displayed more prominent NET infiltrates were the spleen (12/13), kidney (9/13), and lungs (8/13) (Fig 7F, G), while NET detection in the liver was less prevalent (4/13). In 6 cases, we detected NETs in the heart tissue (Fig 7F). The characteristics of these patients are displayed in Table 5. These results confirm that NETs are present in various tissues of patients with severe COVID-19.

**TABLE 5.**
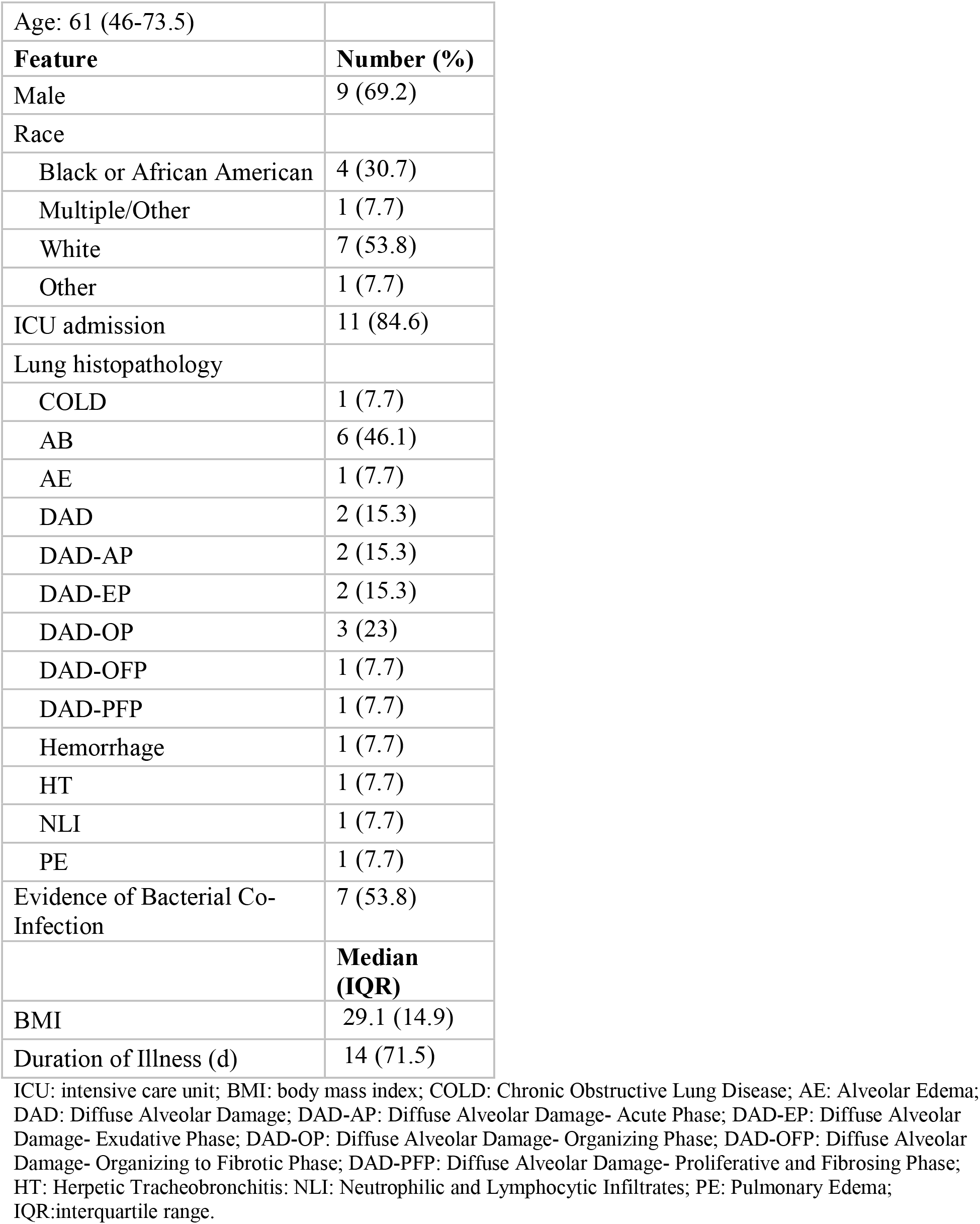
Characteristics of adult patients who died from complications of COVID-19 and whose tissues were used for histologic analysis. Tissue samples cohort (n=13)

|  |  |
| --- | --- |
| Age: 61 (46-73.5) |  |
| **Feature** | **Number (%)** |
| Male | 9 (69.2) |
| Race |  |
| Black or African American | 4 (30.7) |
| Multiple/Other | 1 (7.7) |
| White | 7 (53.8) |
| Other | 1 (7.7) |
| ICU admission | 11 (84.6) |
| Lung histopathology |  |
| COLD | 1 (7.7) |
| AB | 6 (46.1) |
| AE | 1 (7.7) |
| DAD | 2 (15.3) |
| DAD-AP | 2 (15.3) |
| DAD-EP | 2 (15.3) |
| DAD-OP | 3 (23) |
| DAD-OFD | 1 (7.7) |
| DAD-PFD | 1 (7.7) |
| Hemorrhage | 1 (7.7) |
| HT | 1 (7.7) |
| NLI | 1 (7.7) |
| PE | 1 (7.7) |
| Evidence of Bacterial Co-Infection | 7 (53.8) |
|  | **Median (IQR)** |
| BMI | 29.1 (14.9) |
| Duration of Illness (d) | 14 (71.5) |

ICU: intensive care unit; BMI: body mass index; COLD: Chronic Obstructive Lung Disease; AE: Alveolar Edema; DAD: Diffuse Alveolar Damage; DAD-AP: Diffuse Alveolar Damage- Acute Phase; DAD-EP: Diffuse Alveolar Damage- Exudative Phase; DAD-OP: Diffuse Alveolar Damage- Organizing Phase; DAD-OFD: Diffuse Alveolar Damage- Organizing to Fibrotic Phase; DAD-PFD: Diffuse Alveolar Damage- Proliferative and Fibrosing Phase; HT: Herpetic Tracheobronchitis; NLI: Neutrophilic and Lymphocytic Infiltrates; PE: Pulmonary Edema; IQR:interquartile range.

### COVID-19 patients infected with Omicron variant display decreased NET complexes when compared to infections with other variants

During the COVID-19 pandemic, many SARS-CoV-2 variants have emerged (2021). Omicron (B.1.1.529) variant is of particular interest for its rapid spread across the world(Callaway, 2021). Omicron variant harbors 37 mutations in the spike protein(Mannar et al., 2022). Although, Omicron seen to be more contagious, data indicate it is milder than other variants(Nealon and Cowling, 2022). Therefore, we hypothesized that unvaccinated adult patients infected with Omicron variant will display decreased NET levels when compared to those infected with earlier strains (Alpha). Indeed, citH3 and elastase-DNA complexes measured in plasma from unvaccinated Italian COVID-19 patients infected with the Omicron variant were significantly decreased when compared to those that had been infected with the original strain during the first peak of the pandemic (Fig 8 A-B). In addition, males infected with Omicron variant (but not females) displayed significantly lower levels of plasma citH3-DNA complexes when compared to those infected with the original strain (Fig 8C). We found that plasma levels of citH3-DNA complexes were significantly reduced in patients with the Omicron variant that were in critical condition (Fig 8D), those who required ICU admission (Fig 8E), those with hypertension (Fig 8F), and those who required high flow oxygen when compared to those infected with the earlier variant. Overall, these results indicate that different variants of the virus are associated with differences in the levels of circulating NETs and that infection with the Omicron variant is associated with lower levels of these structures when compared to patients infected with earlier strains.

**Figure 8.**
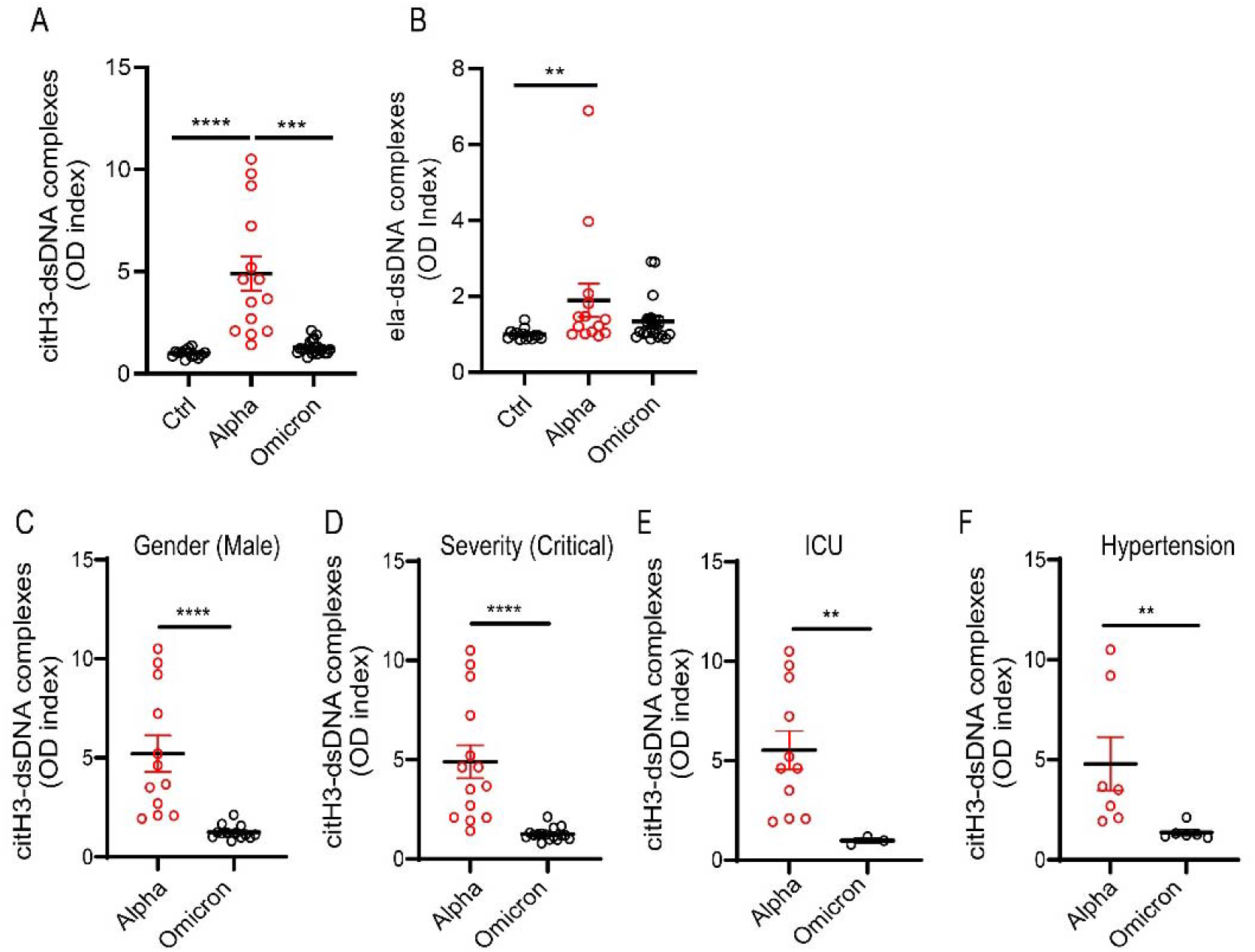
NET remnants are lower in adult unvaccinated patients infected with the Omicron variant. Plasma levels of (**A**) citrullinated histone H3 (citH3) and (**B**) elastase-DNA complexes (ela-dsDNA) were measured in COVID-19 patients infected with SARS-CoV-2 Alpha or Omicron variants (ctrl n= 14, Alpha n=14, Omicron= 21). Kruskal-Wallis analysis was used. (**C**) Male patients infected with the alpha variant of SARS-CoV-2 displayed elevated levels of citH3-DNA complexes (Alpha, n=12, Omicron, n=13). (**D**) Patients with critical severity of disease have lower levels of NETs when infected with Omicron infected patients displayed decreased levels of plasma citH3-DNA complexes (Alpha n= 14, Omicron n= 16). (**E**) COVID-19 patients in the intensive care unit (ICU) displayed decreased elevated levels of citH3-DNA complexes when infected with Omicron variant (Alpha, n=11, Omicron n= 3) (**F**) COVID-19 patients with concomitant hypertension displayed decreased elevated levels of citH3-DNA complexes when infected with Omicron variant (Alpha n= 7, Omicron n= 6), Mann-Whitney was used. Results are the mean +/- SEM. Mann-Whitney was used. **p<0.01, ***p<0.001, ****p<0.0001. OD: optical density

## Discussion

Increasing evidence supports a detrimental role of NETs in COVID-19 pathogenesis. NETs and NET remnants have been detected in adult COVID-19 lung tissues and plasma samples, respectively(Barnes et al., 2020; Zuo et al., 2020). Our findings expand prior work by showing a link between NETs and infection with SARS-CoV-2 in children with MIS-C and CLL, as well as an association between circulating NETs and clinical outcomes in pediatric and adult patients from different geographic locations.

In the present study, we showed that pediatric patients diagnosed with MIS-C and CLL displayed elevated amounts of circulating NETs in serum or plasma and in tissue. Impaired NET degradation was also evident and associated to a variety of factors that include complement activation, presence of anti-NET autoAbs and a natural DNAse1 inhibitor, G-actin. As G-actin levels decreased as the disease improved, this suggests that induction of G-actin may be an acute response phenomenon that occurs in patients with severe COVID-19 who present with impaired NET degradation. Although CLL has remained a controversial entity, the presence of NET remnants across the spectrum of those with confirmed COVID-19 infection in this study supports a SARS-CoV-2 etiology. NET remnants were associated with comorbidities present in pediatric patients and there was geographic variability in these findings. For instance, citH3-DNA levels were elevated in MIS-C samples obtained from Chile, but not from Italy and similar differences in NET degradation capabilities were also observed. While the levels of elastase-DNA were elevated in CLL samples from the United States, they were more evidently elevated in those samples obtained from Italy. While technical factors associated with sample collection and processing at various centers could in theory contribute to these differences, the previous observations that SARS-CoV-2 and various COVID-19-associated host factors can directly induce NET formation in COVID-19(Middleton et al., 2020) support that possibility that environmental or yet unidentified genetic factors, including SARS-CoV-2 variants may modulate how neutrophils respond to the infection and associated inflammatory milieu. It will also be important to assess in the future whether the MIS-C cohort from Chile had stronger associations with autoimmunity features than the cohort of MIS-C patients from Italy, and whether the presence of NETs or NET-associated Abs may show an association with these clinical differences. However, as mentioned in results section, to date none of the patients with MIS-C in Italy or Chile followed in this study have developed persistent autoimmunity.

NETs are dismantled by endogenous DNases that can regulate NET-driven thrombosis.(Barnes et al., 2020; Hakkim et al., 2010; Zuo et al., 2021b) Impairments in NET degradation have been shown to increase endothelial damage,(Carmona-Rivera et al., 2015) associate with thrombus formation,(Middleton et al., 2020; Wolach et al., 2018; Zuo et al., 2021b) organ dysfunction, inflammation and autoimmunity (Knight et al., 2012).We found that pediatric subjects diagnosed with MIS-C or CLL displayed decreased NET degradation that, in most cases, could be restored with exogenous DNase1 in vitro. Genetic and environmental factors contribute to decreased DNase activity or efficiency. Patients harboring mutations in *DNASE1* are at higher risk of developing lupus (Hartl et al., 2021; Yasutomo et al., 2001). However, in the WGS analysis, we did not find genetic drivers of impaired nuclease activity, suggesting that environmental factors, such as differences in microbiome composition, UV light exposure, pollutants, etc., may be involved. Certainly, assessments of larger cohorts of patients will be required to fully exclude genetic drivers of these abnormalities.

Complement activation, presence of natural DNase1 inhibitors, and autoAbs have been reported to contribute to impairments in NET degradation in inflammatory and autoimmune conditions(Hakkim et al., 2010; Leffler et al., 2012; Zuo et al., 2021a). We found that impairments of NET degradation in MIS-C patients were associated with the presence of the natural DNase-1 inhibitor, G-actin, and with enhanced ability for serum C1q to deposit in NETs. In contrast, anti-NET autoAbs were primarily associated with impairments in NET degradation in subjects with CLL. These results highlight the complexity of neutrophil responses following exposure to SARS-CoV-2 and the associations with different disease manifestations. Addition of exogenous DNase1 to MIS-C and CLL samples restored NET degradation capabilities in most samples, suggesting that treatment with exogenous DNases could be of benefit to decrease NETs in COVID-19 patients. Notably, aerosolized DNases treatment have been evaluated in clinical trials as a putative therapy in this disease.

Consistent with previous work, markers of NETs were elevated in adult patients with COVID-19(Zuo et al., 2020). Symptomatic adult COVID-19 samples obtained from Italy during the peak of the pandemic in Europe in early 2020, displayed elevated circulating NETs, while samples from asymptomatic patients did not. While the mechanisms promoting enhanced NET formation in COVID-19 remain to be further determined, these results provide a putative link between NETs and symptomatic COVID-19. COVID-19 associated NET formation may involve other mechanisms beside direct infection of neutrophils by SARS-CoV-2 that explain the lack of NET markers found in infected asymptomatic COVID-19 patients. Furthermore, asymptomatic subjects did not display significant impairments in NET degradation, nor the presence of factors that can interfere with the dismantling of the NETs. In contrast, NETs were elevated in symptomatic COVID-19 patients, and they remained elevated at least 3 months after infection diagnosis. Whether this protracted resolution of NET dysregulation is associated with long-COVID symptoms or other chronic complications remains to be determined and should be the focus of future work. Furthermore, NETs were present in extrapulmonary COVID-19 tissues such as heart, liver, kidney, and spleen. These results may implicate NETs in extrapulmonary manifestations seen in COVID-19 patients, even after clearing the virus.

Several SARS-CoV-2 variants have emerged during COVID-19 pandemic. Omicron is the most recent variant of SARS-CoV-2 reported in late 2021(Callaway, 2021) and was identified as variant of concern because of its rapid spread(Mannar et al., 2022), albeit associated to milder symptoms than other variants(Nealon and Cowling, 2022). NET remnants from adult unvaccinated patients infected with Omicron variant in Italy were significantly reduced when compared to COVID-19 patients infected with the variant of SARS-CoV-2 that affected Italy early in the pandemic. These results could suggest that lower NET formation induced by this variant may explain in part the milder symptomatology. Alternatively, the milder inflammatory phenotype induced by this strain may account for downstream effects on neutrophils, limiting their ability to NET following this infection. Future studies will be needed to further expand the understanding on how different viral strains modulate neutrophil biology, whether some of these differences explain the geographic variation in NET levels and clearance, and the overall implications of these variations in the reported findings.

Various preexisting comorbidities have been associated with higher morbidity and mortality in adult and pediatric subjects that develop COVID-19(Ejaz et al., 2020; Yang et al., 2020). In our study, there was no consistent association between specific comorbidities and levels of NETs in adult and pediatric populations. Pericarditis, kidney damage, neurologic, and psychiatric manifestations are among the clinical manifestations of a subset of the COVID-19 patients(Ejaz et al., 2020; Khaddaj-Mallat et al., 2021; Vinayagam and Sattu, 2020; Yang et al., 2020). While NETs have been detected in lung samples in COVID-19, it remains to be better characterized whether NETs in extrapulmonary tissues during and after COVID-19 in adults and pediatric patients are associated with prognosis, disease manifestations, or development of long COVID. Here, we were able to detect citH4 positive structures in skin from pediatric patients affected by CLL and in autopsy tissues from adult subjects who died from severe COVID-19. Of note, the lung, kidney, and spleen were the organs with most NET detected, followed by the heart. Enhanced NET formation or diminished NET clearance may increase the risk of organ damage, and the presence of NETs in these extrapulmonary tissues may link these structures with other complications associated with COVID-19. As the biological significance of NETs during COVID-19 is still not completely elucidated, future studies should investigate the potentially deleterious role of NETs in extrapulmonary tissues that may explain part of the pathophysiology of severe COVID-19.

Limitations of the study are primarily derived from the heterogeneity of the cohorts. The clinical centers sites did not all collect the same clinical characteristics during the peak of the pandemic, limiting in some cases the comparisons among groups, as mentioned above. Furthermore, at this point, we do not have comprehensive details regarding the development of chronic complications and/or long-COVID ensuing after COVID-19 infection in these cohorts, which would allow us to further elucidate the impact of enhanced NET formation in this condition. It was difficult to assess the impact of specific therapies on NET formation/degradation as detailed information about medication treatment at each center was not available for our analysis. Additionally, we were limited in the collection of samples from patients infected with different SARS-CoV-2 variants. Despite these limitations, we were able to work with heterogenous and geographically diverse cohorts in both pediatric and adult populations.

In summary, our findings support a link between NET dysregulation and pediatric manifestations of COVID-19 in association with specific disease manifestations. Pediatric and adult symptomatic COVID-19 patients displayed impaired NET degradation that did not appear to be genetically driven. These results highlight a putative pathogenic role of NETs and impairments in NET degradation in pediatric and adult subjects affected by COVID-19.

## Materials and Methods

### Sample collection

Subjects recruited in this study fulfilled the Center for Disease Control and Prevention (CDC) Health advisory case definition for MIS-C, CLL and COVID-19. Laboratory confirmation of SARS-CoV-2 infection (positive PCR +/- anti-S/anti-N serology) was assessed. All individuals were enrolled in protocols approved by local Institutional Review Boards (IRBs): Comité Ético Científico Facultad de Medicina Clínica Alemana Universidad del Desarrollo, Santiago, Chile (protocol 2020-41); Comitato Etico Interaziendale A.O.U. Città della Salute e della Scienza di Torino, Turin, Italy (protocol 00282/2020); Ethics Committee of the University of Naples Federico II, Naples, Italy (protocol 158/20); Comitato Etico Provinciale of Brescia, Italy (protocol NP-4000,); University of Milano Bicocca – San Gerardo Hospital, Monza; National Institute of Allergy and Infectious Diseases, National Institutes of Health, Bethesda, MD, USA (protocols NCT04582903, NCT03394053 and NCT03610802); University of Wisconsin-Madison (protocol 2020-0667). The severity of the disease in the pediatric and adult COVID-19 cohorts was defined as follows: 1. asymptomatic, 2. mild, 3. moderate, 4. severe and 5. critical as per the NIH COVD-19 treatment Guidelines (https://files.covid19treatmentguidelines.nih.gov/guidelines/covid19treatmentguidelines.pdf).

For CLL samples obtained from the University of Wisconsin-Madison, patients were prospectively enrolled and consented after the start of the COVID-19 pandemic. Those with prior history of seasonal pernio were included due to speculation for a shared genetic susceptibility with pandemic-associated pernio with other circulating viruses as a possible trigger. Affected skin biopsy specimens were obtained from those requiring tissue diagnosis for standard of care. These were formalin-fixed, paraffin-embedded and sectioned prior to histopathologic diagnosis of CLL. Laboratory assessments were performed at day 0, at 6-8 weeks, and at 6 months. Samples from healthy age, gender and sex-matched ctrls without history of COVID-19 symptoms or PCR-confirmed infection were used for serological studies.

### Post-mortem tissue collection

Autopsies were performed and tissues were collected as previously described(Huang et al., 2021b) in the National Cancer Institute’s Laboratory of Pathology at the NIH Clinical Center, coordinated by the NIH COVID-19 Autopsy Consortium and following consent of the legal next of kin.

### Whole Genome Sequencing (WGS)

Genomic DNAs were extracted from patients’ whole blood by an automated nucleic acid sample preparation instrument (Qiagen QIAsymphony SP) using the QIAsymphony DNA Midi Kit. DNA samples were then quantified using a fluorescence dye-based assay (PicoGreen dsDNA reagent) by a microplate reader (Molecular Devices SpectraMax Gemini XS). WGS libraries were generated from fragmented DNA using the Illumina TruSeq DNA PCR-Free HT Library Preparation Kit with minor modifications for automation (Hamilton STAR Liquid Handling System). Sequencing libraries were quantified using the KAPA qPCR Quantification Kit (Roche Light Cycler 480 Instrument II) and combined as 24-plex pools after normalization and sequencing on an Illumina NovaSeq 6000 using a S4 Reagent Kit (300 cycles) using 151+8+8+151 cycle run parameters. Primary sequencing data was demuxed using the Illumina HAS2.2 pipeline and sample-level quality control for base quality, coverage, duplicates, and contamination was conducted. All sequencing date were then processed with Burrows–Wheeler Aligner (BWA) and the Genome Analysis Toolkit (GATK) best-practice pipeline for alignment and variant calling. Whole genome association data were deposited at dbGaP under accession number phs002245.v1.p1

### NET-complexes ELISA

A 96-well plate was coated with rabbit polyclonal anti-citH3 (Abcam ab5103) or anti-neutrophil elastase (Calbiochem, 481001) at 2.5 ug/mL in PBS overnight at 4°C. Wells were blocked in blocking buffer (1% BSA in PBS) at RT for 1 h. Plasma or serum were diluted 1: 100 in blocking buffer and incubated overnight at 4°C. After washing three times with washing buffer (0.05% Tween in PBS), samples were incubated with mouse monoclonal anti-double stranded DNA antibody (EMD Millipore, Clone BV16-13, MAB030) at 1:100 in blocking buffer for 1 h at RT. Plate was washed three times and incubated with goat anti-mouse conjugated HRP antibody (1:10,000) (Bio-Rad, 1721012) for 1h at RT. Wells were washed five times with 0.05% Tween in PBS followed by the addition of 100 uL of TMB substrate (Sigma Aldrich) and 50 uL of 0.16M sulfuric acid stop solution (Sigma Aldrich). The absorbance was measured at 450 nm on an ELISA plate reader.

### NET degradation assay

Control neutrophils were resuspended in un-supplemented RPMI (1×10^6^ cells/mL) and were stimulated with PMA (Sigma) (500ng/mL). A hundred microliters/ well were plated in a 96-well black plate and incubated for 4 h at 37°C to induce NETs. Following stimulation, formed NETs were treated with 5% serum from either healthy ctrls or patients for 16 h at 37°C. Wells were stained with 0.2 uM Sytox green (ThermoFisher) for 5 min. Plate was read using a microplate reader (Synergy HT, Bio-Tek). Results are presented as relative fluorescence units (RFU). Wells were visualized to corroborate the presence or the absence of NETs using the ZOE microscope.

### Detection of NETs in tissue specimens

Parafilm embedded samples were processed as previously described.(Carmona-Rivera et al., 2017)

### Immunofluorescence

Neutrophils were fixed in 4% paraformaldehyde in PBS overnight at 4°C, washed, and blocked with 0.2% porcine gelatin (Sigma, St. Louise, MO) for 30 min, then incubated with primary antibody (anti-C1q or patient serum) for 1 h in a humid chamber at 37°C. Coverslips were then washed three times and incubated 30 min with secondary antibody at 37°C. Nuclei were counterstained with 1:1000 Hoechst at RT. After washing three more times, coverslips were mounted on glass slides using Prolong-gold solution (Invitrogen). Images were acquired on a Zeiss LSM 780 confocal microscope.

### Anti-DNase1, anti-DNase1L3 and NET Ab detection

A 96-well plate was coated with 2.5 ug/mL of PMA-generated NETs, recombinant DNase1 (Abcam) or DNase1L3 (MyBiosource) in PBS overnight at 4°C. Plate was blocked with 1% BSA for 1 h at RT. One microgram of total protein from control or patient skin samples or diluted (1:200) sera were incubated overnight at 4°C. Plate was washed 4 times with 0.05% PBS-Tween (PBS-T). Horseradish conjugated-anti-human IgG secondary antibody (1:5,000; Sigma) was incubated for 1 h at RT, followed by 5 washes with PBS-T. Plate was developed in the presence of TMB and read at 450 nm using a microplate reader (Synergy HT, Bio-Tek). Results are presented as optical density (OD) index (ratio of the OD in the patient serum to the mean OD in healthy control serum).

### G-actin ELISA

Serum G-actin levels were detected using commercially available G-Actin ELISA kit (MyBioSource) following the manufacturer’s instructions.

### Statistical analysis

Data was processed using R version 4.0 and the tidyverse package. All figures and associated statistics were performed using GraphPad Prism Version 8.1.1 (La Jolla, CA). Mann-Whitney U test was used for pair-wise comparisons. One-way analysis of variance (ANOVA) Kruskal-Wallis test (Dunn’s multiple comparison test) was used to compare parameters among groups. Pearson correlation was used for all non-categorical statistics. All analyses were considered statistically significant at p < 0.05.

## Data Availability

All data produced in the present study work are contained in the manuscript

## Author contribution

C. Carmona-Rivera designed and performed the experiments, analyzed the data, and wrote the manuscript. T. Markowitz, D.R. Claybaugh participated in statistical analysis of clinical samples. H.C. Su, Y. Zhang, A. Oler and C. Dalgard performed WGS analysis. K. Dobbs, R. Castagnoli, H. Kenney, D. Draper, M. Truong, O.M. Delmonte, F.Licciardi, U. Ramenghi, N. Crescenzio, L. Imberti, A. Sottini, V. Quaresima, C. Fiorini, V. Discepolo, A. Lo Vecchio, L. Pierri, A. Catzola, M. C., A. Biondi, P. Bonfanti, M.C., Poli Harlowe, Y. Espinoza, C. Astudillo, E. Rey-Jurado, C. Vial, J. de la Cruz, R. Gonzalez, C. Pinera, J. W. Mays, A. Ng, A. Platt, B. Drolet, J. Moon, E.W. Cowen, S. Weber, M. Magliocco and L.D. Notarangelo collected and provided clinical specimens and clinical data, reviewed manuscript, and provided scientific input. K. Dobbs, S. Weber, H. Su, D. Chertow, L.D. Notarangelo provided advice and helpful suggestions on project design. J.W. Mays, A. Ng, A. Platt, B. Drolet, S. Hewitt, L.M. Arkin, D.S. Chertow provided tissue specimen and clinical data. M.J. Kaplan planned the project, supervised the work and wrote the manuscript.

## Acknowledgments

Supported by the Intramural Research Program at NIAMS/ (ZIAAR041199 to MJK), NIAID (ZIA AI001270-01 to LDN and ZIAAI001265 to HCS), NIDCR (ZIADE000747 to JM),the Wisconsin Partnership Program 2020 Strategic COVID-19 Grant (to LA), FONDECYT 11181222 (to MCP), ANID 0999 (to MCV and MCP), and Regione Lombardia. We thank Smilee Samuel, Jason Barnett and Sandhya Xirasagar for Labkey infrastructure, Helen Matthews for logistical help, Justin Lack for biostatistical support.

The contents of this publication do not necessarily reflect the views, opinions or policies of the USUHS or the Department of Defense.

