## Supplementary Tables for "Multicenter analysis of neutrophil extracellular trap dysregulation in adult and pediatric COVID-19"

Supplementary Materials

**SUPPLEMENTARY TABLE 1: WHOLE GENOME SEQUENCING ANALYSIS OF NON DEGRADERS AND DEGRADERS IN PEDIATRIC SAMPLES**

Excel sheet

**SUPPLEMENTARY TABLE 2: CORRELATIONS BETWEEN ELASTASE: DNA COMPLEXES AND CLINICAL VARIABLES IN SERUM SAMPLES**

| **FEATURE** | **VARIABLE (N);**  ***MEDIAN*** | **VARIABLE (N);**  **MEDIAN** | **P Value** |
| --- | --- | --- | --- |
| Sex | F (71); 1.750 | M (176); 1.435 | 0.04329 |
| Age | <60 (120); 1.669 | >60 (127); 1.421 | 0.05880 |
| Severity | Mild (20); 6.740 | Moderate (18); 1.392 | 0.00086 |
| Severity | Mild (20); 6.740 | Severe (41); 1.473 | 2.16E-07 |
| Severity | Mild (20); | Critical (156); 1.405 | 9.56E-09 |
| Oxygen needs | None (14); 1.737 | Low Flow (83); 1.341 | 0.27880 |
| Oxygen needs | None (14); 1.737 | High Flow (22); 1.720 | 0.85834 |
| Oxygen needs | None (14); 1.737 | Intubation (31); 1.489 | 0.71304 |
| Oxygen needs | None (14); 1.737 | Tracheostomy (27); 1.288 | 0.24261 |
| Pneumonia | Absent (30); 5.596 | Present (203); 1.406 | 4.82E-07 |
| Renal Insufficiency | Absent (227); 1.495 | Present (6); 1.423 | 0.64097 |
| Death | Alive (200); 1.620 | Deceased (43); 1.202 | 0.04125 |
| Need for ICU | ICU (73); 1.404 | non-ICU (174); 1.620 | 0.06302 |
| Comorbidities | Absent (183); 1.406 | Present (45); 1.746 | 0.02078 |
| CVD | Absent (178); 1.529 | Present (50); 1.278 | 0.11870 |
| Hypertension | Absent (137); 1.523 | Present (91); 1.357 | 0.52106 |
| Diabetes Mellitus | Absent (185); 1.495 | Present (43); 1.480 | 0.93352 |
| CHF | Absent (209); 1.489 | Present (19); 1.480 | 0.96089 |
| CKD | Absent (186); 1.500 | Present (42); 1.415 | 0.43488 |
| Obesity | Absent (207); 1.489 | Present (21); 1.398 | 0.95432 |
| Hematologic Malignancy | Absent (177); 1.518 | Present (51); 1.404 | 0.27880 |
| Solid Malignancy | Absent (204); 1.500 | Present (24); 1.412 | 0.68980 |
| Neurologic/Psychiatric Disease | *Absent* (182); 1.488 | Present (46); 1.485 | 0.51216 |
| Autoimmunity | Absent (180); 1.460 | Present (48); 1.718 | 0.15354 |
| Chronic Respiratory Disease | Absent (206); 1.492 | Present (22); 1.477 | 0.42819 |
| Liver Disease | Absent (210); 1.485 | Present (18); 1.611 | 0.99851 |
| Solid Organ Transplant | Absent (211); 1.505 | Present (17); 1.357 | 0.71938 |

**SUPPLEMENTARY TABLE 3: CORRELATIONS BETWEEN CITRULLINATED HISTONE H3: DNA COMPLEXES AND CLINICAL VARIABLES IN PLASMA SAMPLES**

| **FEATURE** | **VARIABLE (N);**  ***MEDIAN*** | **VARIABLE (N);**  **MEDIAN** | **P Value** |
| --- | --- | --- | --- |
| Sex | F (17); *2.008* | M (73); *1.461* | 0.70290 |
| Age | <60 (36); *1.343* | ≥60 (54); *1.756* | 0.55043 |
| Severity | Mild (2); *2.988* | Moderate (3); *3.714* | 0.80000 |
| Severity | Mild (2); *2.988* | Severe (14); *2.060* | 0.81667 |
| Severity | Mild (2); *2.988* | Critical (65); *1.448* | 0.24581 |
| Oxygen needs | None (1); *4.215* | Low Flow (22); *2.032* | 0.34783 |
| Oxygen needs | None (1); *4.215* | High Flow (5);*3.254* | 0.66667 |
| Oxygen needs | None (1); *4.215* | Intubation (14); *0.902* | 0.26667 |
| Oxygen needs | None (1); *4.215* | Tracheostomy (21); *1.013* | 0.18182 |
| Pneumonia | Absent (7); *2.934* | Present (77); *1.461* | 0.04833 |
| Renal Insufficiency | Absent (81); *1.569* | Present (3); *2.932* | 0.51518 |
| Death | Alive (74); *1.458* | Deceased (12); *1.783* | 0.67630 |
| Need for ICU | ICU (41); *1.013* | non-ICU (48); *2.260* | 0.01554 |
| Comorbidities | Absent (77); *1.752* | Present (6); *0.738* | 0.00949 |
| CVD | Absent (56); *1.343* | Present (27); *2.067* | 0.03368 |
| Hypertension | Absent (44); *1.076* | Present (39); *2.016* | 0.13392 |
| Diabetes Mellitus | Absent (62); *1.489* | Present (21); *1.752* | 0.97911 |
| CHF | Absent (73); *1.569* | Present (10); *2.016* | 0.47991 |
| CKD | Absent (62); *1.19* | Present (21); *2.481* | 0.04486 |
| Obesity | Absent (74); 1.646 | Present (9); *1.53* | 0.74175 |
| Hematologic Malignancy | Absent (55); *1.752* | Present (28); *1.265* | 0.95008 |
| Solid Malignancy | Absent (73); *1.569* | Present (10); *1.764* | 0.81745 |
| Neurologic/Psychiatric Disease | *Absent (62); 1.549* | Present (21); *1.752* | 0.62620 |
| Autoimmunity | Absent (58); *1.424* | Present (25); 2*.008* | 0.39607 |
| Chronic Respiratory Disease | Absent (74); *1.646* | Present (9); 1*.059* | 0.35237 |
| Liver Disease | Absent (75); *1.448* | Present (8); *3.320* | 0.01713 |
| Solid Organ Transplant | Absent (73); *1.387* | Present (10); *3.151* | 0.00803 |

ICU: intensive care unit; CVD: cardiovascular disease; CHF: congestive heart failure; CKD: chronic kidney disease.

**SUPPLEMENTARY TABLE 4: WHOLE GENOME SEQUENCING ANALYSIS OF NON DEGRADERS AND DEGRADERS IN ADULT SAMPLES**

Excel sheet
